# Nothing but hot air? – On the molecular ballistic analysis of backspatter generated by and the hazard potential of blank guns

**DOI:** 10.1101/2020.12.21.20248446

**Authors:** Jan Euteneuer, Annica Gosch, Cornelius Courts

**Affiliations:** Institute of Forensic Medicine, University Medical Center Schleswig-Holstein, Kiel, Germany

**Keywords:** molecular ballistics, backspatter, ballistic model, blank cartridge guns, blank guns, wound channel

## Abstract

Blank cartridge guns are prevalent especially in countries with laws restricting access to conventional firearms and it is a common misconception that these weapons are harmless and only used as toys or for intimidation. However, although their harming potential is well documented by numerous reports of accidents, suicides, and homicides, a systematic molecular biological investigation of traces generated by shots from blank cartridges at biological targets has not been done so far.

Herein, we investigate the occurrence and analyzability of backspatter generated by shots of different types of blank cartridge guns firing different types of blank ammunition at ballistic gelatin model cubes doped with human blood and radiological contrast agent soaked into a spongious matrix and covered with three different variants of skin simulants.

All skin simulants were penetrated and backspatter was created in 100% of the shots in amounts sufficient for forensic STR typing that resulted in correct identification of the respective blood donor. Visible backspatter was documented on the muzzle and/or inside the barrel in all cases, and in 75% of cases also on the outer surfaces and on the shooter’s hand(s).Wound cavities were measured and ranged between 1 and 4.5 cm in depth.

Discussing our findings, we provide recommendations for finding, recovering and analyzing trace material from blank guns; and presenting a comprehensive overview of the pertinent literature on injuries inflicted by blank guns we emphasize the considerable hazard potential of these devices.

## Introduction

Gun ownership is widespread around the world and incidents of gunshot related injuries and deaths are well documented [1] and in some countries so prevalent that gun violence had to be declared as a “public health crisis” [2]. Blank cartridge guns, or blank firing guns, on the other hand, and due to the lack of a projectile, are regarded by many as less hazardous to be used as toys or non-lethal devices for self-defense. Hence, they are freely available in many countries for adults and can be purchased without any need for registration, and with no proper regulation in place. They are most prevalent in countries where access to conventional firearms is limited and regulations for gun ownership and gun use are restrictive. Example for this are Turkey, where blank guns are cheap and blank gun-related fatalities common (e.g. [3]), and Germany, where in 2020 the police union (GdP, “Gewerkschaft der Polizei”) estimated that about 15 million blank cartridge guns are circulating in the public. This is nearly three times the number of weapons and weapon parts registered at the German national weapon register (NWR, “Nationales Waffenregister”) [4]. Meanwhile the number of “small gun licenses”, which are required to carry a blank gun in public, is about 670,000, thus representing only 4.5% of the estimated blank guns owned in Germany [4].

These numbers indicate the need for and relevance of ballistic research on blank cartridge guns and ammunition and their hazard potential. However, although numerous case reports are available, experimental studies focusing on blank guns are limited. Apparently, their ability to severely injure or even kill is highly underrated, even though comments on their potential hazard including warnings against misuse had been published as far back as 1865 [5]. About one hundred years later, more case reports began to emerge and Shepard among others reported incidents involving wounds caused by blank cartridges and corroborated the warnings issued before, calling the designation “blank” a “misnomer” [6]. Tausch et al. also demonstrated the danger posed by blank ammunition to human bodies and advocated for stricter legal regulations in Germany already in 1974 [7]. More recent studies employing ballistic gelatin models with focus on wound morphology were published for instance by Schyma & Schyma [8] and Pircher et al. [9]. However, systematic research involving molecular ballistic analyses of biological traces created by blank cartridge gun shots at biological targets has not been performed so far with the exception of a single study analyzing traces of backspatter inside 20 guns after fatal contact shots to the head, that included a .380 revolver described as a blank cartridge gun from which sufficient trace material for STR typing could be retrieved [10]. Therefore, more research is necessary, as, given that lack of a projectile, regular ballistic knowledge is transferable only to a limited degree to blank cartridge gunshot phenomena.

In this study, we apply “molecular ballistics” (the molecular biological analysis of biological traces originating from gunshots at biological targets) to blank cartridge gunshots using different types of weapons and ammunition at variations of ballistic gelatin models with skin simulant, simulating a torso. From the results we intend to infer the DNA profile of “the victim” and assess the deposition of trace material. For this purpose, we aim to investigate whether ‘backspatter’ [11], the biological material which is propelled from the entrance wound back towards the shooter and which has been shown to persist inside of firearms [12], is reliably created by guns emitting only a gas jet without a projectile. The generation of backspatter by conventional firearms has been known for a long time [13] and its molecular ballistic analyzability has been proven not only in real cases e.g. of multiple homicide [14], but also experimentally [15, 16]. In the present study, we are first to evaluate the possibility of STR profiling from backspatter traces generated by blank cartridge gunshots. We consider difficulties of handling and offer recommendations for finding and sampling biological traces from blank cartridge guns. Also, we evaluate the hazard potential of blank guns based on wound channel analysis and by discussing the literature and anecdotal evidence pertaining to the destructive potential of a blank cartridge gas jet.

## Materials and Methods

### Blood collection and sample mixtures

The venous blood employed in the sample mixtures was drawn by venipuncture and collected in sterile K3 EDTA S-Monovettes® (Sarstedt, Germany). It was donated by two informed and consenting adult volunteers, neither of whom were involved in the loading or shooting process at the shooting site, nor sample collection, nor weapon cleaning. EDTA blood of each donor was mixed with the CT-contrast agent Barilux® (Sanochemia Diagnostics, Germany) in a ratio of 1:1 to obtain a “double-contrast” mixture as described recently [15], representing a modification of a previously established multi-component contrast mixture [16].

### Preparation of the ballistic models

The ballistic models used in this study were intended to roughly emulate a human torso. Hence, they were based on the “reference cube” introduced by Schyma et al. [17] and were constructed as three slightly different variants: (1) A single layer of chamois leather of 10 cm x 10 cm was glued onto the bottom of a polypropylene box, on top of which a spongious matrix, soaked with about 20 mL of “double-contrast” mix and tightly wrapped in cling film, was glued as well; (2) the same set-up as in (1) but with two layers of chamois leather glued together; and (3) a single layer of chamois leather, but with the wrapped “double-contrast” sponge additionally sealed into an evacuated vacuum bag. These variants were chosen to represent possible physiological variation of differences in skin thickness [18] and mechanical properties [19] between individuals and different locations on the torso. Chamois Leather with a thickness of about 1-2 mm was chosen as a skin simulant following a recommendation for ballistic test shootings by a ballistics expert of the German Federal Criminal Police Office (personal communication).

The polypropylene boxes were then filled with type III ballistic gelatin (Honeywell Fluka™, Germany) prepared at a 10% concentration following Fackler’s instructions [20] and were subsequently stored for about 36 hours at 4 °C. Prior to shooting, the gelatin block was taken out of the box (the leather would stick to the gelatin and peel from the bottom despite the glue) and cut into blocks of about 12 cm x 14 cm x 11 cm.

### Experimental shooting setup

The experimental shooting was conducted at the designated shooting area on the premises of the State Office of Criminal Investigation of Schleswig-Holstein (LKA-SH) in Kiel. The gelatin blocks were placed upon a table onto a stiff base of foam rubber padding, to enable proper gelatin expansion without giving too much leeway for movement [21]. All shots were contact shots aiming at the center of the leather skin simulant that were executed by one of the authors freehand and without mechanical stabilization of the weapon to simulate realistic shooting conditions. To avoid contamination by the shooter, forensic-grade protective gear used in the Institute of Forensic Medicine Kiel, Germany, Standard Earloop Facemasks (3M Health Care, Germany) and Micro-Touch® nitrile examination gloves (Ansell, Belgium) were worn when and changed before each shooting.

Two currently freely available and commonly encountered 9 mm blank cartridge guns were used in this study: one pistol (Ekol Firat Compact, Voltran Silah Sana, Turkey) and one revolver (Zoraki R1 2.5”, ATAK Arms Industry, Turkey). In addition, one 8 mm blank pistol (Reck Commander 8 mm, UMAREX GmbH, Germany), which is not sold anymore but still common as a collector’s item, was retrieved from the stock of the LKA-SH. The weapons and different kinds of ammunition employed herein are listed in Table 1, including one black powder and one non C.I.P^1^. -listed ammunition.

After each shot, the gelatin blocks were wrapped in plastic foil and transferred back into the plastic boxes for further evaluation.

### Trace documentation, sampling, and weapon cleaning

To obtain an impression of the respective backspatter distribution, the weapons, as well as the hands of the shooter and any additional noticeable traces were photographed after each shot.

Sampling was conducted using DNA-free forensic nylon swabs (4N6 FLOQ Swabs Genetics, Copan Flock Technologies, Italy) and by applying a modified double swab technique [22], where a single swab is moistened on one half with 20 µL HPLC gradient grade water (Th. Geyer GmbH & Co. KG, Germany), while the other half remains dry. Backspatter samples were collected from distinct sampling locations termed A – D (in cases without visible traces, the entire location was swabbed): the outer frame / slide of the weapon (A), the muzzle and surrounding frame at the front (B), and the outer surfaces of the barrel, which is exposed by pulling back the slide (C, pistols only). Swabbing the inside of the barrel (D) cannot be properly conducted using the above mentioned standard forensic nylon swabs as it is hindered by the blockage attached within the barrel of commercially available blank guns; hence, in this study, thin stripes of medical cellulose were inserted into the barrel and shoved past the blockage as far as possible with the help of a pipette tip to collect any traces of backspatter (see Supp. Fig. 1 E, 6 B). The cellulose stripes had been treated with UV light for 24 hours prior to the shooting and had been experimentally tested to be free of amplifiable human DNA. Afterwards the pistols were disassembled (the revolver had no detachable pieces), and the parts examined for further traces.

Prior to each shot after sampling and documentation, all surfaces were thoroughly cleaned mechanically with Kimtech Science Precision Wipes (Kimberly-Clark, USA) and chemically using Roti®-Nucleic Acid-free (Carl Roth GmbH, Germany) and distilled water. The inside of the barrel was rinsed along the blockage with both liquids and scrubbed with Roti®-Nucleic Acid-free-soaked pipe cleaners. The weapons were then reassembled and subsequently blow-dried by the shooter with compressed air. Finally, and immediately before the next shot, negative controls were taken from all sampling locations of the weapon.

### DNA extraction, quantification and STR profiling

DNA extraction from all samples was performed using the PrepFiler® Forensic DNA Extraction Kit (Thermo Fisher Scientific, USA) according to the manufacturer’s recommendations, with the exception of additionally employing NucleoSpin Forensic Filters (Macherey-Nagel, Germany) in the lysis step, yielding an elution volume of 50 µL.

The DNA concentration was determined by quantitative PCR (qPCR) using the PowerQuant® system (Promega, Wisconsin, USA) on an Applied Biosystems™ 7500 fast Realtime PCR System (Thermo Fisher Scientific). Quantification was performed in duplicates following the manufacturer’s instructions, but as a modified approach with 2 µL DNA sample in a reduced reaction volume of 10 µL. This approach had been validated and accredited for routine analysis in our laboratory. DNA degradation and PCR inhibition values, as put out by the PowerQuant® analysis tool, were analyzed as well to evaluate the DNA quality from samples created with different blank ammunitions and taken from different sampling locations.

For one sample from each backspatter event and for negative samples exhibiting an amount of quantifiable autosomal DNA above 0.4 pg/µL (lab-internally validated threshold below which no relevant amplification of human STR alleles is to be expected), STR multiplex-PCR was performed to investigate possible contaminations using the NGM Detect™ PCR Amplification Kit (Thermo Fisher Scientific) according to the manufacturer’s protocol on an Applied Biosystems™ GeneAmp PCR System 9700 (Thermo Fisher Scientific) and with an optimal input amount of 0.5 ng DNA, if possible. PCR products were separated and detected on an Applied Biosystems™ 3500 Genetic Analyzer (Thermo Fisher Scientific) Data was analysed with the GeneMapper ID-X software version 1.5 (Thermo Fisher Scientific) and mixed profiles evaluated with the LRmix Studio software version 2.1.5 [23, 24].

### Ballistic Model and wound channel evaluation

The gelatin cube models were photographed, and the extent of the entrance “wound” on the leather skin simulant documented. Afterwards the wound channel was dissected from the gelatin block and serially cut to about 0.5 cm thick slices. A scanner (MP C306, Ricoh, Canada) was used to create images of the slices (600 dpi). Those images were further evaluated with ImageJ 1.52d (NIH, USA), first qualitatively, and second, where possible, quantitatively by applying the polygon method [25, 26], where the end of the tears of the wound cavity are connected, creating a polygon whose circumference reflects the extent of the damage inflicted by the gas jet on the gelatin.

### Literature research

A thorough and comprehensive browsing of available research literature in German and English was conducted to identify forensically relevant studies, reviews and case reports concerning blank firing guns and ammunition. Articles of interest were identified using the Pubmed.gov (U.S. National Library of Medicine) search engine with search terms as “blank guns AND ballistics”, “blank guns AND forensic” etc., as well as by perusing further references listed in these publications. Articles with purely medical focus on treatment were excluded, as well as articles which were referenced in publications, but could not be found and validated online. The collection can be found in Supp. Table 2 and is presented as a sortable Excel table.

## Results and discussion

### General appearance of backspatter

The goal of this study was to systematically assess whether backspatter in forensically relevant amounts is generated in a reproducible manner by shots from and propelled back onto and into blank guns. To this aim, we employed different kinds and types of weapons and ammunitions as well as ballistic models with three variants of skin simulant.

We demonstrated that backspatter traces containing DNA of sufficient amount and quality for STR profiling could be detected on the inner and/or outer surfaces of every weapon following each shot that penetrated through the contrast mixture bag into the gelatin block (examples shown in Fig. 1). All but one shot penetrated the respective skin simulant, the exception being one shot from the revolver firing “Walther 9 mm R.K.” ammunition at a double layer of skin simulant. This is proof that the gas jet alone as produced by blank guns is sufficient to reproducibly create the energy and cause the correspondent wound ballistic effects (e.g. a temporal wound cavity) that are required for backspatter generation. Additionally, the shooter’s hand demonstrated visible traces of backspatter after 75% of the shots (Table 2, Fig. 1 I), and after 33% of shots, different amounts of backspatter were found on the PPE, goggles and/or face mask of the shooter (Table 2, Supp. Fig. 5 D).

### Distribution of backspatter traces and molecular ballistic analysis

A summary of the results is presented in Table 2. Sampling was performed in a non-quantitative manner (as no replicates of weapon-ammunition-model combinations were performed), i.e. in cases where extensive amounts of backspatter occurred, only one sample swab was taken. Otherwise, the entire sampling location was swabbed. The major part of backspatter generated by contact shots was expected to consolidate at and around the muzzle (sampling location B) and inside the barrel (sampling location D). Indeed, visible traces at the muzzle could be detected after 100% of the shots (Fig 1 A-C, Supp. Fig. 1-12) and all corresponding swab samples yielded DNA concentrations sufficient to perform STR profiling (between 9.2 pg/µL and 1.6 ng/µL). Traces within the barrel could not as easily be detected however, as the barrel blockage stretches through nearly the entire barrel short of a few millimeters of the muzzle, rendering photo documentation difficult (e.g. Supp Fug. 7 B – 9 B), even with the use of additional lamps. Despite these difficulties, backspatter inside the barrel was detectable by visual inspection in 75% of the shots. Endoscopic examination, even though an endoscope could not be completely inserted into the barrel of a blank gun, nevertheless might be of help to detect possible traces, as it does with regular handguns [27], by providing an illuminated camera view from the muzzle inside the barrel with adjustable zoom and viewing angle. Our method of inserting cellulose stripes into the barrel (Supp. Fig. 1 E, 6 B) proved to be cumbersome but effective for collecting sufficient backspattered material from 100% of the shots. DNA from the stripes could be readily extracted applying a standard procedure with concentrations ranging between 6.6 pg/µL and 4.5 ng/µL. In cases when traces are hardly or not visible at all, however, alternative sampling techniques may be appropriate, as the stripes cannot be shoved through the entire length of the barrels. Barrel blockages come in different shapes and sizes depending on the gun’s type, model and manufacturer, and the sampling technique needs to be adapted to the weapon at hand. In any case, the space between blockage and inner surface of the barrel will not be wider than a few millimeters (see e.g. Fig. 1 C), too narrow to fit in neither standard forensic swabs nor even the finest swabs available to us (4N6FLOQ Crime Scene Divisible Swabs (Copan)). Future studies could assess medical swabs with mini tips for their applicability, e.g. pediatric, urethral, or specimen collection swabs like the “FLOQSwabs Sterile Ultra Thin Minitip” (Copan) or the “Sterile Mini-tip Rayon/Polyester Swabs” (Puritan, USA), both with ca. 2 mm tip diameter. However, an alternative or additional approach to collect biological material from inside the barrel could be to carefully rinse the barrel with water or buffer and collect the flowthrough. When cleaning the weapons after each shot, we observed that a considerable amount of our “double-contrast” mixture persisted inside the barrel and could be washed out (Fig. 1 G). Such an approach could be tested with dried samples and anticoagulant-free blood in future work.

Backspatter on the frame, the outer surface of the slide and other exterior parts (sampling location A) was documented after 75% of shots (Table 2). Three cases were free of visible traces, and quantification of the DNA extracted from samples recovered from these locations resulted in concentrations of 0.1, 0.3 and 0.5 pg/µL, i.e. slightly below or above our lab-internal threshold of 0.4 pg/µL for successful STR profiling (Table 2). Still, all three weapons bore clearly visible and analyzable backspatter traces inside the barrel and at the muzzle, respectively. Lacking high-speed recordings of the process, possible explanations for the variability in trace distribution are speculative but include twitching of the shooter’s hand, variation of extent and pattern of the rupture of the leather skin simulant at the entry site, chaotic distribution of the muzzle gas inside and between the model components (“double-contrast”-bag, gelatin and skin simulant), as well as variations of the combustion of the propellant, and any combination of those.

In criminal cases, the standard procedure for trace recovery from a gun should require its complete disassembly and sampling of inner surfaces [14]. With blank firing guns mostly built up of few easily detachable pieces, this can be done accordingly. While the revolver in this study had no detachable parts, both pistol models did, yet the smaller parts, e.g. the recoil spring, did hardly bear any visible traces in our sample set (only found after one pistol shot, see Supp. Fig. 5 E), unlike as had been described in previous work with regular hand guns [15]. The exception from this was the inner surface of the detachable slide (Fig. 1 J), which together with the outer surface of the barrel was subsumed as sampling location C. Sampling those areas yielded DNA concentrations above 0.4 pg/µL in 85% of cases, with a maximum of 666.6 pg/µL.

Due to the generally large amounts of recoverable backspatter, DNA profiling was straightforward and resulted in full STR profiles that correctly identified the respective blood donor i.e. the simulated victim in 100% of the samples. Only one profile exhibited three additional minor drop-in alleles, yet all below 2.5% average peak height.

### Evaluation of negative controls

After each shot, firearms were thoroughly cleaned to remove remaining traces from previous shots. Out of 42 negative controls, however, 12 (29%) yielded DNA concentrations above 0.4 pg/µL, between 1 and 48.5 pg/µL (Supp. Table 1). Of those, 10 originated either from sampling location B or D, i.e. the muzzle area and inside the barrel, where most of the backspatter was detected and effective cleaning had proven to be most difficult due to the barrel blockage. In 7 of 12 cases, STR profiling identified the respective blood donor of the previous shot, indicating that even our thorough and extensive cleaning procedure was insufficient. While this, on the one hand, demonstrates the need to optimize DNA removal when cleaning blank guns, it portends, on the other hand, an opportunity in real cases to retrieve traces from inside the barrel even from guns that had been thoroughly cleaned. In 3 of 12 cases, a mixed profile of two donors with distinct major and minor components were found: in one sample, this was most likely due to contamination, as the minor contributor (all peaks <15% average peak height) had performed the DNA extraction. The other two samples originated from the same shot; there, the minor contributor was the blood donor for the model that had been shot before with the very same weapon. Therefore, minimal traces appeared to have remained inside the weapon even after two rounds of extensive cleaning, although cross contamination during the sample processing cannot be excluded as well. As no contamination was detected at the regular samples collected after the shot, the large amounts of backspatter seem to have ‘quenched’ or masked the minuscule remains of that respective donor. From the last 2 of 12 negative controls, one was a mixed profile with small peaks not clearly attributable to any donor or involved person, and the other represented the STR profile of the shooter, probably due to a contamination while air-drying the weapon after cleaning. There were, however, no STR signals attributable to the shooter found in the following or any subsequent shot from this weapon.

### Recommendations for the sampling of blank firing guns

Taking together the findings described so far, it can be constituted that contact shots delivered by different types of blank guns firing different kinds of ammunition at different types of ballistic models reproducibly generate sufficient amounts of backspatter to allow for molecular ballistic analysis. This observation will highly likely hold true also for real biological targets. It therefore needs to be discussed, where exactly to find and how to collect such traces, especially considering the structure of the barrel.

As seen in our samples, a variety of locations can be reached by the backspatter traces, thus, as with real handguns, the entire weapon should be examined carefully, and all parts disassembled if possible. Sampling should be performed starting with the outside parts (beginning with visible traces on the frame, trigger guard and so on) followed by the inside parts. The muzzle is the most obvious outside part for backspatter to be recovered from. If the gun is cleaned and free of visible traces, all parts, especially inner surfaces of detachable parts or the outer surface of the barrel, should still be examined and sampled as well. Pooling of samples will in general increase the chance of recovering sufficient material for DNA analysis [10]. Finally, the barrel’s inside as a prominent source of backspatter that is well shielded against the environment, should be sampled using techniques and tools that are matched to the structure of the barrel blockage. The technique applied herein using cotton stripes may be appropriate, but can certainly be adapted, improved, and optimized. One approach, especially with cleaned guns, could be to rinse the barrel directly with lysis buffer for DNA extraction and pool the flow-through with sampled material on cotton stripes for a combined lysis step. In our hands, cleaning the barrel even with chemicals specifically dedicated to DNA removal, was not always sufficient to remove all remains, which may be exploited for evidence collection in real cases.

### Evaluation of the hazard potential of blank guns

#### Examination of the entrance wound and wound cavity

The contrast mixture used in this study comprised venous blood and an X-ray contrast agent to facilitate radiological wound channel evaluation via CT analysis as described before [15, 28]. However, the wound channels produced by the blank gun shots proved to be small and mostly filled with gunshot residues (GSR), barring meaningful results from CT analysis which was hence discarded. Alternatively, the extent of the destruction in the gelatin was quantified applying the polygon method to the analysis of 0.5 cm cut slices and add the resulting values to a sum representing the total damage (Table 2). Alternative methods based on crack length were considered, but as the wound channel frequently did not exhibit a clear center lacking a proper bullet path, the measurement of crack lengths would have been inaccurate.

Little data is available on the range of energy transfer of blank cartridge ammunition. Still, it appeared plausible to expect that the depth of the cavity would in our experimental setup depend on the thickness of the skin simulant when shot with the same blank gun and ammunition, while its appearance (shape and position) would be attributable to the kind of ammunition, albeit with considerable variation, as seen in previous studies using gelatin e.g. by Schyma et al. and Pircher et al. [8, 9]. Despite employing a different model setup, wound morphologies and depths were comparable to these studies. An expectable correlation of wound depth and skin simulant thickness could not be confirmed with our sample number though (all depths of the wound channels listed in Table 2). Representative examples of and all wound cavity morphologies are displayed in Fig.2, and Supp. Fig. 14, respectively. Their shape was essentially tubular or conical, with varying width and distinct blackening from the GSR in all cases. In two cases, both produced with the revolver firing “Walther 9 mm R.K.” ammunition, the wound cavity was not sufficiently deep to be cut out or was even nondetectable at all. Given that the “double-contrast” mix bag had been penetrated and backspatter had indeed been created, a minimum gas jet length of 1 cm may be deduced, roughly equaling the combined thickness of skin simulant and bag. The maximal length of a cavity in our study resulted from a shot with black powder ammunition with about 4.5 cm, hence as well matching the above mentioned findings of Schyma et al. and Pircher et al., but also substantiating a considerable wounding potential when inflicted to real biological targets i.e. living organisms. A notable difference was observed when comparing ammunition based on nitrocellulose and black powder employed in two shots: In addition to the greater depth and more pronounced gelatin destruction as inferred by the polygon perimeter values, black powder ammunition also produced a distinct yellow and grey shaped ring around the wound cavity and discoloration throughout the entire cavities. This phenomenon had not been described in the literature before, but we suggest that this may be an effect of a chemical or thermal reaction between the burning black powder and the gelatin.

At every block, the skin simulant was slightly ballooning convexly towards the outside due to the backspattered gas (best seen at Supp. Fig. 14 F & H). The gas jet entry sites were lacerated by multiple, mostly 3 to 4 tears (Fig 1 D-E), which rather resembled blank cartridge gunshots to the head, e.g. as shown in [29]. The wounds were accompanied by, albeit small, substance losses of the skin simulant in 75% of cases, while in the remaining cases (two with double layers of chamois leather and one with vacuum bag) the ‘wounds edges’ of the skin simulant were perfectly adaptable (Supp. Fig. 13). This is in contrast to the findings of Pircher et al. [9], who reported roundish skin defects produced by their experimental shots at their pig skin-gelatin model. The different characteristics of chamois leather regarding tensile strength [30] as well as the “double-contrast” mixture bag being positioned underneath the skin simulant, thus offering a softer space for the leather to spread into, might be a possible explanation for this.

#### Further examples and observations of the damage potential from blank guns

The harming potential has been illustrated not only by our experiments or the referenced studies, but by numerous reported incidents of accidents, homicides, suicides and further experiments, which are listed in a considerable literature collection presented in the supplements, which we compiled to our best knowledge and effort (Supp. Table 2).

Also, in addition to our data presented so far and taking into account the abstract nature of experiments with gelatin model setups, we believe that it may be useful and instructive to present some more quite illustrative if only anecdotal evidence of the drastic effect of and hence danger posed by blank firing guns of different kinds. Firstly, we fired the Ekol pistol and Skullfire ammunition at an empty, anatomically realistic skull model made from polyurethane (SYNBONE, Switzerland) that simulates the mechanical properties of human bone and whose forensic use has been described elsewhere [28, 15]. Several shots penetrated the skull simulant that had been covered with a double layer of chamois leather as skin simulant, creating considerable defects in the model (Supp. Fig 15). We could not reliably reproduce this effect with skull models that were filled with gelatin, still these findings do correspond to what has been reported from real cases and suggest that shots to the head with blank guns can be experimentally reconstructed and repeated.

Secondly, the massive destructive potential (contact shot) and potential thermal damage (near distance shot) of blank gun shots is illustrated in Supp. Fig. 16: an uncooked whole chicken (intended for consumption) was shot at with a blank pistol, applying a contact shot and shot from close distance. The results demonstrate the potential severe harm to skin and muscle tissue of biological targets. Thirdly, a further illustration of the energy transfer and damage inflicted by ‘mere’ muzzle gas is presented in Supp. Fig. 17, where the gas jets produced by blank gun fire are shown to punch through a several mm wooden board.

In addition to these examples and the reports of real cases in Suppl. Table 2, it should be emphasized that, due to the free and unregulated availability of blank cartridge guns, an unknown proportion – probably even the majority – of cases involving these weapons are not reported in literature or included in freely available official statistics. It remains one of the most conspicuous legal discrepancies of arms regulations and concerns for public health and safety, that access to devices with time and again proven capacity to inflict potentially deadly harm is scarcely controlled and regulated at all.

## Data Availability

not applicable

## Conclusions

Herein, we present the first systematic demonstration that backspatter is reproducibly generated by shots from different types of blank cartridge guns firing different kinds of ammunition at different variations of ballistic models doped with a source of biological trace material. Application of molecular ballistics has been shown to be suitable and is therefore advisable for the analysis of backspatter trace material that consolidates and persists on outer and inner surfaces of the weapon.

Blank guns are intended to mimic the appearance of regular live arms but are constructed in different manner, with the barrel blockage being the most prominent feature. It obstructs the barrel in a manner that considerably impedes trace collection and cleaning, but also creates the opportunity in real crime cases to retrieve minute amounts of trace material even after the weapon has been cleaned.

The severe harming potential of the gas jet from blank ammunition, regardless if nitrocellulose or black powder, could be confirmed and is in concordance with prior experiments and case reports involving blank firing guns.

## Declarations

### Ethics approval

This study was performed in line with the principles of the Declaration of Helsinki. The study design and experimental procedures were approved by the ethics committees of the University Hospital of Bonn and the University Medical Center of Schleswig-Holstein.

### Availability of data and material

Raw and additional data are available upon request.

### Code availability

Not applicable.

## Acknowledgements

We, again, are very grateful to the Landeskriminalamt Schleswig-Holstein for facilitating and friendly help with the shooting session on their premises. Special thanks go to Jan “Punk’s not dead” Rohwer for providing us with high quality blood samples and to Franziskarrr “Risikobegegnung” Siefer for her help during shooting sessions. We also thank Phil Cachée (Sachverständigenbüro Cachée, Berlin, Germany) for providing us with shocking chicken footage. The expert technical assistance of Kathi “Iiieh, baaah!” Pöhls is also gratefully acknowledged. Finally, we thank the Deutsche Forschungsgemeinschaft (DFG) for funding this project (CO 992/7-1).

## Footnotes

1 C.I.P.: Commission internationale permanente pour l’épreuve des armes à feu portatives, engl. “*Permanent International Commission for the Proof of Small Arms”*

