## Supplementary material for "Nothing but hot air? – On the molecular ballistic analysis of backspatter generated by and the hazard potential of blank guns": Tables

| **Table 1. Weapons and ammunition** | | | | | |
| --- | --- | --- | --- | --- | --- |
| **Weapon** | **Manufacturer** | **Ammunition** | **Material** | **Propellant** | **Manufacturer** |
| Pistol EKOL Firat Compact | Voltran Silah Sana (Turkey) |  |  |  |  |
|  |  | Skullfire  9 mm P.A.K. | Steel | NC | Pobjeda Technology (Bosnia and Herzegovina) |
|  |  | Özkursan  9 mm P.A.  (Not C.I.P. listed) | Nickel | NC | Özkursan (Turkey) |
| Revolver Zoraki R1 2.5” | ATAK Arms Industry (Turkey) |  |  |  |  |
|  |  | Walther  9 mm  R.K / R.B. | Brass | NC | Carl Walther GmbH (Germany) |
|  |  | Geco  9 mm R Knall / .380 R Blanc | Brass | NC | RUAG Ammotec GmbH (Germany) |
|  |  | Center Fire  Blank Cartridges  .380 / 9 mm | Brass | BP | Dynamit Nobel AG (Germany) |
| Pistol Reck Commander PTB 238 | UMAREX GmbH (Germany) |  |  |  |  |
|  |  | Walther  8 mm K. | Brass | NC | Carl Walther GmbH (Germany) |
| NC: Nitrocellulose, BP: Black powder, P.A.K.: Pistole Automatik Knall (Pistol automatic bang), P.A.: Pistol automatic, R.K.: Revolver Knall (Revolver bang), K.: Knall (bang), C.I.P.: Commission internationale permanente pour l'épreuve des armes à feu portatives | | | | | |

| **Table 2. Summary of backspatter trace evaluation and wound cavity assessment** | | | | | | | | | | | | | | | |
| --- | --- | --- | --- | --- | --- | --- | --- | --- | --- | --- | --- | --- | --- | --- | --- |
| **Weapon** | **Ammunition** | **Model** | **Visible Backspatter** | | | | | | **Molecular Ballistics Results** | | | | | **Wound Cavity** | |
| **Manufacturer**  **type - caliber** | **Manufacturer**  **Propellant** | **Setup** | **on Shooter** | | **at Sampling Location** | | | | **DNA Quantity at Sampling Location** | | | | **Full STR Profile of Donor** | **Depth [mm]** | **Polygon Perimeter Sum [cm]** |
|  |  |  | **Hand(s)** | **Body** | **A** | **B** | **C** | **D** | **A** | **B** | **C** | **D** |  |  |  |
| Zoraki  R – 9 mm | Walther  NC | 1 | X | - | X | X | n.a. | X | +++ | +++ | n.a. | +++ | X | 11 | n.a. |
| Zoraki  R – 9 mm | Walther  NC | 3 | - | - | X | X | n.a. | - | ++ | ++ | n.a. | + | X | 10 | n.a. |
| Zoraki  R – 9 mm | Geco  NC | 2 | X | - | X | X | n.a. | - | +++ | +++ | n.a. | ++ | X | 24 | 8.45 |
| Zoraki  R – 9 mm | Nobel  BP | 1 | X | X | X | X | n.a. | X | +++ | +++ | n.a. | +++ | X | 45 | 24.33 |
| Zoraki  R – 9 mm | Nobel  BP | 3 | X | - | X | X | n.a. | X | ++ | ++ | n.a. | + | X | 40 | 32.30 |
| EKOL  P – 9 mm | Skullfire  NC | 1 | X | X | X | X | X | X | + | ++ | ++ | +++ | X | 25 | 14.37 |
| EKOL  P – 9 mm | Skullfire  NC | 3 | - | - | - | X | - | X | ++ | ++ | + | +++ | X | 21 | 3.10 |
| EKOL  P – 9 mm | Skullfire  NC | 2 | X | - | X | X | - | X | ++ | + | - | ++ | X | 14 | 10.26 |
| EKOL  P – 9 mm | Özkursan  NC | 2 | X | X | X | X | X | X | ++ | ++ | +++ | +++ | X | 27 | 8.47 |
| EKOL  P – 9 mm | Özkursan  NC | 3 | X | X | X | X | X | X | +++ | +++ | ++ | ++ | X | 29 | 20.13 |
| Reck  P – 8 mm | Walther  NC | 1 | - | - | - | X | X | X | - | +++ | +++ | +++ | X | 25 | 7.69 |
| Reck  P – 8 mm | Walther  NC | 2 | X | - | - | X | X | X | - | + | ++ | ++ |  | 30 | 13.10 |
| For a detailed description of weapons and ammunition, see Table 1. R: Revolver, P: Pistol, NC: Nitrocellulose, BP: Black Powder, Model Setups: 1: single layer chamois leather 2: double layer chamois leather 3: single layer chamois leather + vacuum bag, n.a.: not applicable, Weapon/sampling locations: A: Frame/Slide, B: Muzzle, C: Outer Barrel Surface, D: Inside Barrel, +++: > 0.5 ng/µL, ++: 0.05 - 0.5 ng/µL, +: 0.0004 - 0.05 ng/µL, -: < 0.0004 ng/µL | | | | | | | | | | | | | | | |
