## Supplementary material for "Nothing but hot air? – On the molecular ballistic analysis of backspatter generated by and the hazard potential of blank guns": Figure Legends

**Figure 1**

**A** Backspatter traces on Ekol pistol, after shot with Özkursan 9 mm ammunition **B** Backspatter traces on Zoraki revolver, after shot with Nobel black powder 9 mm ammunition **C** Backspatter traces on Reck pistol, after shot with Walther 8 mm ammunition **D** Gelatin block with double layer of chamois leather, after shot with Özkursan 9 mm ammunition **E** Gelatin block with single layer of chamois leather, after shot with Nobel black powder 9 mm ammunition **F** Gelatin block with single layer of chamois leather, after shot with Walther 8 mm ammunition **G** Flowthrough of barrel (see main text for context) **H** Backspatter traces on the revolver’s recoil shield **I** Backspatter on shooting hand **J** Backspatter traces on the slide of the Ekol pistol

**Figure 2**

Wound channels in gelatin blocks after shots with **A** Ekol pistol, Özkursan 9mm nitrocellulose ammunition, to single layer chamois leather with vacuum bag, **B** Zoraki revolver, Nobel black powder 9 mm ammunition, to single layer chamois leather with vacuum bag, **C** Reck pistol, Walther 9 mm nitrocellulose ammunition, to single layer of chamois leather
