## Supplementary material for "Nothing but hot air? – On the molecular ballistic analysis of backspatter generated by and the hazard potential of blank guns": Suppl. Figure Legends

**Supplementary Figure Legends**

**Supp. Fig. 1 Backspatter documentation**

Weapon: Ekol Firat pistol

Ammunition: Skullfire 9 mm

Model Setup: Single layer chamois leather

A: View outer surfaces, B: View muzzle/barrel, C: Hands of shooter, D: Process of cleaning the barrel using a pipe cleaner, E: Sampling technique of backspatter traces within the barrel using cellulose stripes

**Supp. Fig. 2 Backspatter documentation**

Weapon: Ekol Firat pistol

Ammunition: Skullfire 9 mm

Model Setup: Double layer chamois leather

A: View Outer surfaces, B: View muzzle/barrel, C: Hands of shooter, D: Detachable parts of pistol

**Supp. Fig. 3 Backspatter documentation**

Weapon: Ekol Firat pistol

Ammunition: Skullfire 9 mm

Model Setup: Single layer chamois leather + vacuum bag

A: View outer surfaces, B: View muzzle/barrel, C: Hands of shooter, D: Flowthrough with biological traces after rinsing the barrel

**Supp. Fig. 4 Backspatter documentation**

Weapon: Ekol Firat pistol

Ammunition: Özkursan 9 mm

Model Setup: Double layer chamois leather

A: View outer surfaces, B: View muzzle/barrel, C: Hands of shooter, D: Detachable slide with traces on inner surface

**Supp. Fig. 5 Backspatter documentation**

Weapon: Ekol Firat pistol

Ammunition: Özkursan 9 mm

Model Setup: Single layer chamois leather + vacuum bag

A: View outer surfaces, B: View muzzle/barrel, C: Hands of shooter, D: Backspatter traces on shooter’s clothes and mask, E: Traces on small detachable parts of pistol

**Supp. Fig. 6 Backspatter documentation**

Weapon: Zoraki R1 Revolver

Ammunition: Walther 9 mm

Model Setup: Single layer chamois leather

A: View outer surfaces, B: View muzzle/barrel, C: Hands of shooter, D: Flowthrough with biological traces after rinsing the barrel

**Supp. Fig. 7 Backspatter documentation**

Weapon: Zoraki R1 Revolver

Ammunition: Geco 9 mm

Model Setup: Double layer chamois leather

A: View outer surfaces, B: View muzzle/barrel, C: Hands of shooter, D: Backspatter traces visible on cylinder and trigger guard

**Supp. Fig. 8 Backspatter documentation**

Weapon: Zoraki R1 Revolver

Ammunition: Walther 9 mm

Model Setup: Single layer chamois leather + vacuum bag

A: View outer surfaces, B: View muzzle/barrel, C: Hands of shooter, D: View from top

**Supp. Fig. 9 Backspatter documentation**

Weapon: Zoraki R1 Revolver

Ammunition: Nobel 9 mm Black Powder

Model Setup: Single layer chamois leather

A: View outer surfaces, B: View muzzle/barrel, C: Hands of shooter, D: Traces collected from cartridge chamber in cylinder

**Supp. Fig. 10 Backspatter documentation**

Weapon: Zoraki R1 Revolver

Ammunition: Nobel 9 mm Black Powder

Model Setup: Single layer chamois leather + vacuum bag

A: View outer surfaces, B: View muzzle/barrel, C: Hands of shooter, D: Backspatter traces visible on cylinder and trigger guard

**Supp. Fig. 11 Backspatter documentation**

Weapon: Reck Commander Pistol

Ammunition: Walther 8 mm

Model Setup: Single layer chamois leather

A: View outer surfaces, B: View muzzle/barrel, C: Hands of shooter, D: Minimal traces at pipe cleaner from inside the barrel

**Supp. Fig. 12 Backspatter documentation**

Weapon: Reck Commander Pistol

Ammunition: Walther 8 mm

Model Setup: Double layer chamois leather

A: View outer surfaces, B: View muzzle/barrel, C: Hands of shooter

**Supp. Fig. 13**

Entrance Site at gelatin blocks after shots

A: Weapon: Ekol Firat pistol

Ammunition: Skullfire 9 mm

Model Setup: Single layer chamois leather

B: Weapon: Ekol Firat pistol

Ammunition: Skullfire 9 mm

Model Setup: Double layer chamois leather

C: Weapon: Ekol Firat pistol

Ammunition: Skullfire 9 mm

Model Setup: Single layer chamois leather + vacuum bag

D: Weapon: Ekol Firat pistol

Ammunition: Özkursan 9 mm

Model Setup: Double layer chamois leather

E: Weapon: Ekol Firat pistol

Ammunition: Özkursan 9 mm

Model Setup: Single layer chamois leather + vacuum bag

F: Weapon: Zoraki R1 Revolver

Ammunition: Walther 9 mm

Model Setup: Single layer chamois leather

G: Weapon: Zoraki R1 Revolver

Ammunition: Geco 9 mm

Model Setup: Double layer chamois leather

H: Weapon: Zoraki R1 Revolver

Ammunition: Walther 9 mm

Model Setup: Single layer chamois leather + vacuum bag

I: Weapon: Zoraki R1 Revolver

Ammunition: Nobel 9 mm Black Powder

Model Setup: Single layer chamois leather

J: Weapon: Zoraki R1 Revolver

Ammunition: Nobel 9 mm Black Powder

Model Setup: Single layer chamois leather + vacuum bag

K: Weapon: Reck Commander Pistol

Ammunition: Walther 8 mm

Model Setup: Single layer chamois leather

L: Weapon: Reck Commander Pistol

Ammunition: Walther 8 mm

Model Setup: Double layer chamois leather

**Supp. Fig. 14**

Wound Cavity Morphologies with side view and 0.5 cm cut slices (scanned with 600 dpi)

A: Weapon: Ekol Firat pistol

Ammunition: Skullfire 9 mm

Model Setup: Single layer chamois leather

B: Weapon: Ekol Firat pistol

Ammunition: Skullfire 9 mm

Model Setup: Double layer chamois leather

C: Weapon: Ekol Firat pistol

Ammunition: Skullfire 9 mm

Model Setup: Single layer chamois leather + vacuum bag

D: Weapon: Ekol Firat pistol

Ammunition: Özkursan 9 mm

Model Setup: Double layer chamois leather

E: Weapon: Ekol Firat pistol

Ammunition: Özkursan 9 mm

Model Setup: Single layer chamois leather + vacuum bag

F: Weapon: Zoraki R1 Revolver

Ammunition: Walther 9 mm

Model Setup: Single layer chamois leather

Wound cavity not deep enough to cut

G: Weapon: Zoraki R1 Revolver

Ammunition: Geco 9 mm

Model Setup: Double layer chamois leather

H: Weapon: Zoraki R1 Revolver

Ammunition: Walther 9 mm

Model Setup: Single layer chamois leather + vacuum bag

Wound cavity not deep enough to cut

I: Weapon: Zoraki R1 Revolver

Ammunition: Nobel 9 mm Black Powder

Model Setup: Single layer chamois leather

J: Weapon: Zoraki R1 Revolver

Ammunition: Nobel 9 mm Black Powder

Model Setup: Single layer chamois leather + vacuum bag

K: Weapon: Reck Commander Pistol

Ammunition: Walther 8 mm

Model Setup: Single layer chamois leather

L: Weapon: Reck Commander Pistol

Ammunition: Walther 8 mm

Model Setup: Double layer chamois leather

**Supp. Fig. 15**

Skull model (SYNBONE, Switzerland) shot with Ekol First pistol and Skullfire ammunition to a glued-on skin simulant (double layer chamois leather) causing large defects.

A Right side of skull, B Left side of skull, both defects at the area between temporal and parietal bone.

**Supp. Fig. 16**

Shots at raw frying chicken with Zoraki 9 mm P.A.K. model 917 pistol

A, B Contact shot at chicken breast

C, D Shot from ca. 5 cm distance at chicken leg

E-H Shot from ca. 5 cm at chicken leg over time

(photos by courtesy of P. Cachée (Sachverständigenbüro Cachée, Berlin, Germany))

**Supp. Fig. 17**

Contact shots to wooden board

A Contact shot, caliber 9 mm P.A. bang, SRS Pistol Walther / Umarex, Model P22, PTB 778

B Contact shot, caliber 9 mm P.A. bang with pepper irritant

(photos by courtesy of the Landeskriminalamt Schleswig-Holstein, Kiel, Germany)
