## Supplementary material for "Nothing but hot air? – On the molecular ballistic analysis of backspatter generated by and the hazard potential of blank guns": Suppl. Table 1S

**Supplementary Tables**

| **Supp. Table 1. Summary and evaluation of negative controls with quantifiable DNA amounts** | | | | | | | |
| --- | --- | --- | --- | --- | --- | --- | --- |
| **Weapon** | **Ammunition** | **Model** | **Negative Control** | **Molecular Ballistics Results** | | | **Evaluation** |
| **Manufacturer**  **type - caliber** | **Manufacturer**  **Propellant** | **Setup** | **Sampling Location** | **DNA quantity [ng/µL]** | **Full STR systems from donor** | **Extra alleles** |  |
| Zoraki, R – 9 mm | Walther, NC | 1 | A | 0.0034 | **14/16** | 23 | Extra alleles minor contributor (<15% average peak height), attributable to author doing sample processing. Likely contamination. |
| Zoraki, R – 9 mm | Walther, NC | 1 | B | 0.0275 | **16/16** | - | Unsuccessful cleaning. |
| Zoraki, R – 9 mm | Walther, NC | 1 | D | 0.0062 | **16/16** | 2 | Minor drop-in alleles. Unsuccessful cleaning. |
| Zoraki, R – 9 mm | Walther, NC | 3 | B | 0.0325 | **16/16** | - | Unsuccessful cleaning. |
| Zoraki, R – 9 mm | Geco, NC | 2 | A | 0.0458 | **16/16** | - | Unsuccessful cleaning. |
| Zoraki, R – 9 mm | Geco, NC | 2 | B | 0.0061 | **16/16** | 2 | Minor drop-in alleles. Unsuccessful cleaning. |
| EKOL, P – 9 mm | Skullfire, NC | 2 | B | 0.0010 | 0/16 | **28** | Full STR profile from shooter. Contamination. |
| EKOL, P – 9 mm | Özkursan, NC | 2 | B | 0.0026 | **16/16** | 1 | Minor drop-in alleles. Unsuccessful cleaning. |
| Zoraki, R – 9 mm | Nobel, BP | 1 | A | 0.0287 | **16/16** | 4 | Minor drop-in alleles. Unsuccessful cleaning. |
| Zoraki, R – 9 mm | Nobel, BP | 1 | B | 0.0115 | **16/16** | 15 | Extra alleles minor contributor (<15% average peak height), attributable to donor of block shot before. Likely unsuccessful cleaning. |
| Zoraki, R – 9 mm | Nobel, BP | 1 | D | 0.0010 |  | 26 | Weak profile (63 – 385 RFU) with small peaks, not attributable to one person. Unsuccessful cleaning. |
| Reck, P – 8 mm | Walther, NC | 1 | B | 0.0020 | **9/16** | 6 | All alleles, also extra alleles attributable to donor. Unsuccessful cleaning. |
| For a detailed description of weapons and ammunition, see Table 1. R: Revolver, P: Pistol, NC: Nitrocellulose, BP: Black Powder, Model Setups: 1: single layer chamois leather 2: double layer chamois leather 3: single layer chamois leather + vacuum bag, n.a.: not applicable, Weapon/sampling locations: A: Frame/Slide, B: Muzzle, C: Outer Barrel Surface, D: Inside Barrel. **Bold** in STR systems: major contributor | | | | | | | |
