## Supplementary material for "Nothing but hot air? – On the molecular ballistic analysis of backspatter generated by and the hazard potential of blank guns": Suppl. Figures

### Slide 1
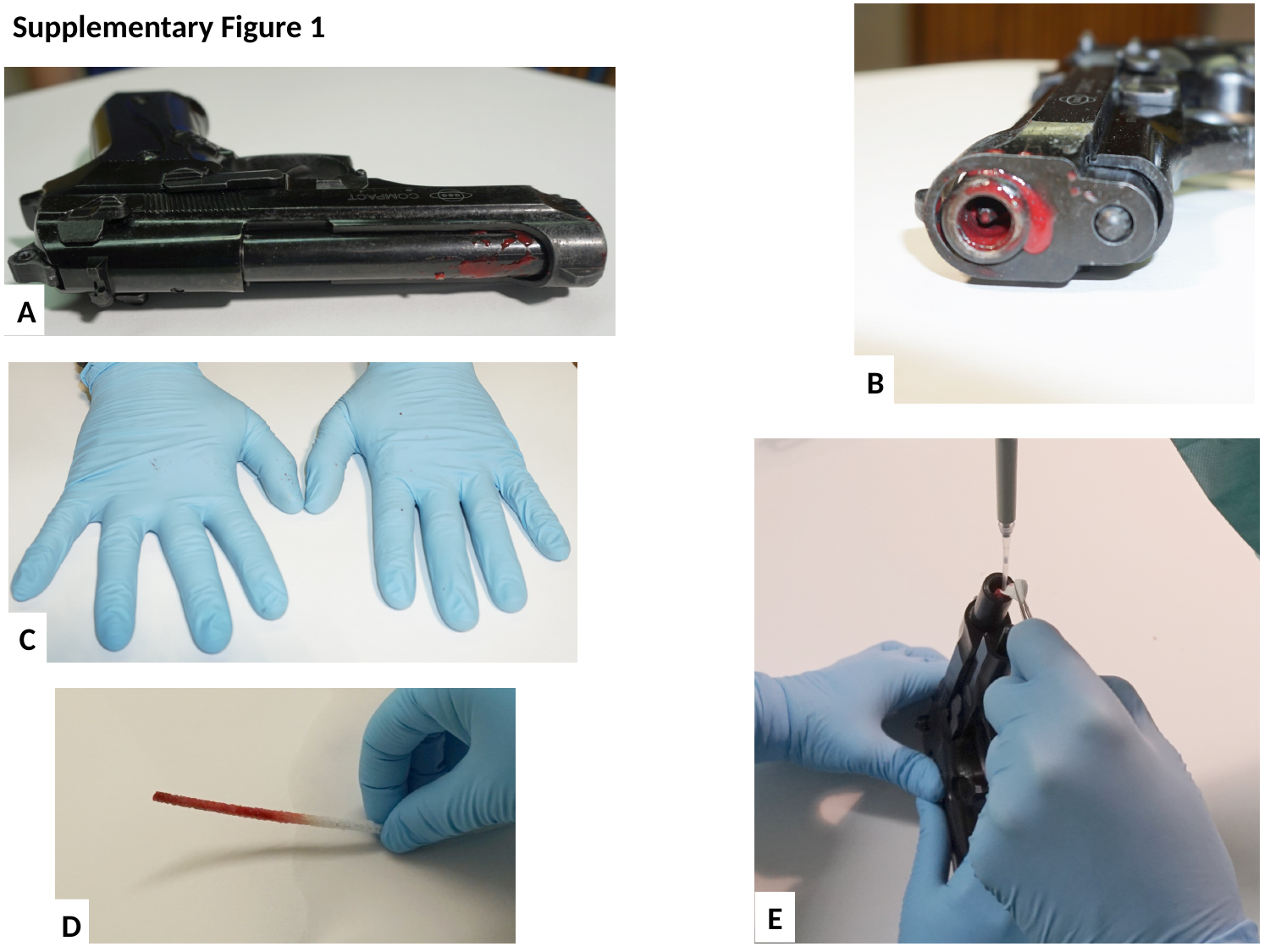

Supplementary Figure 1
A
B
C
E
D

### Slide 2
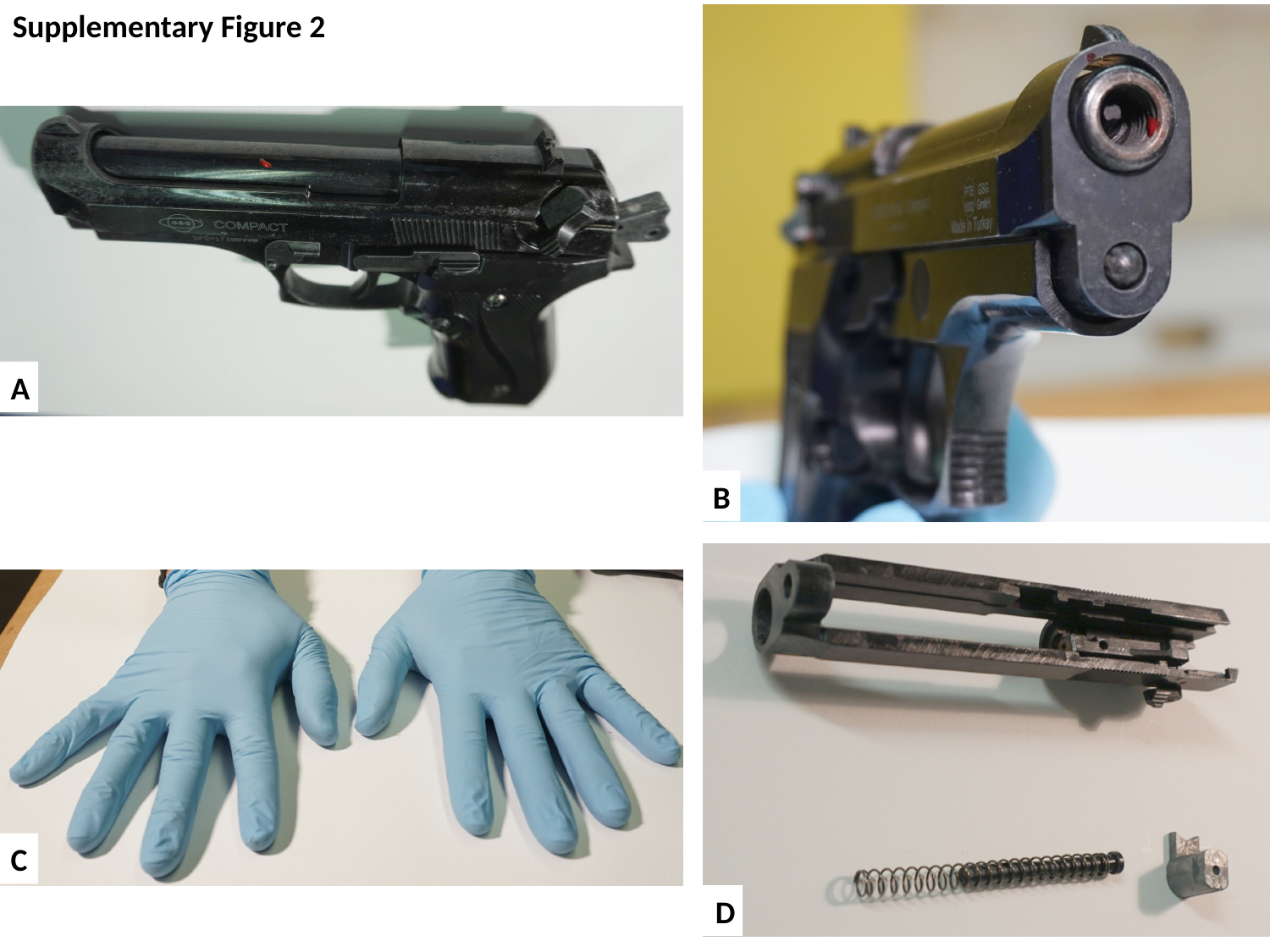

Supplementary Figure 2
A
B
C
D

### Slide 3
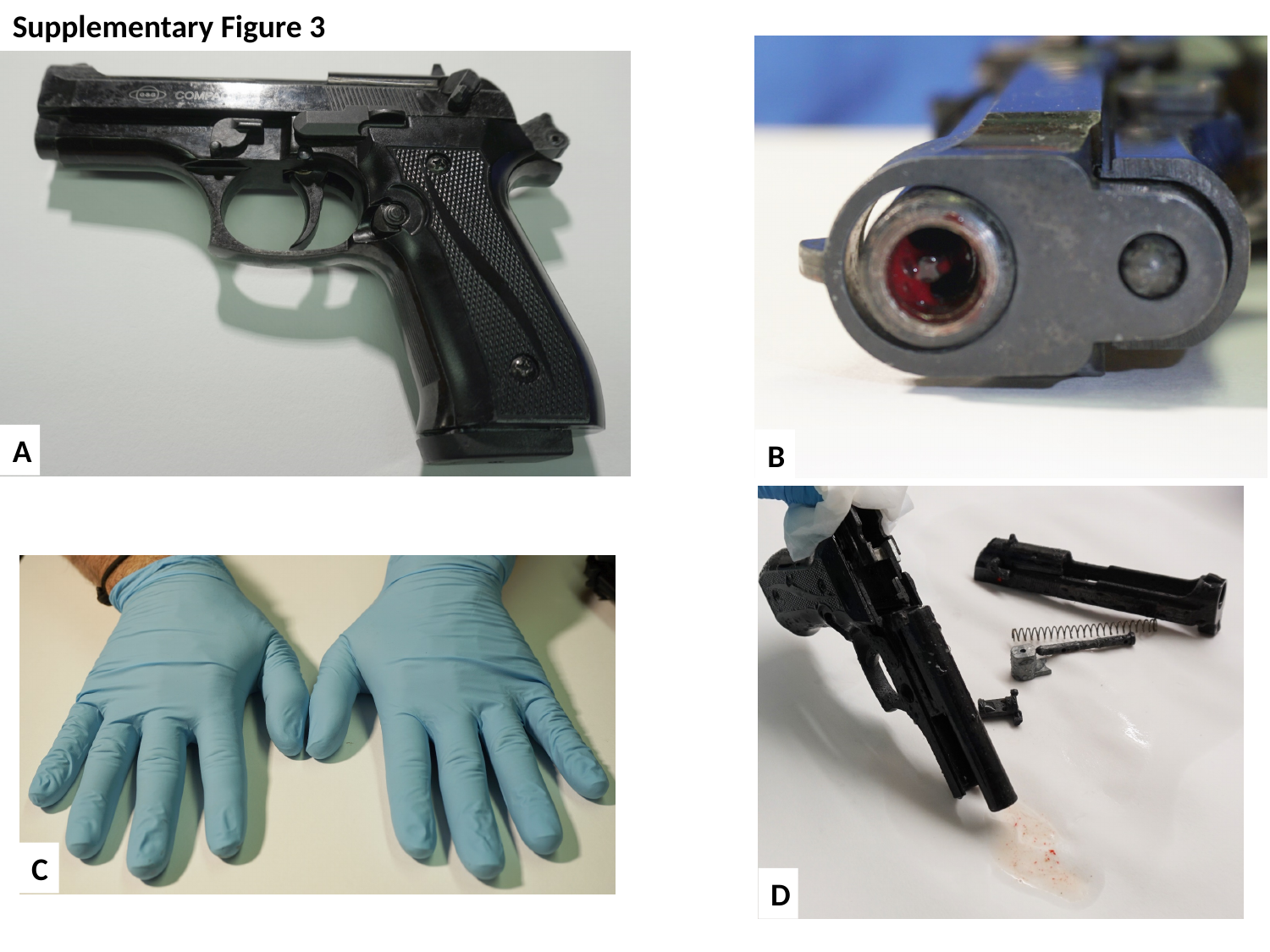

Supplementary Figure 3
A
B
C
D

### Slide 4
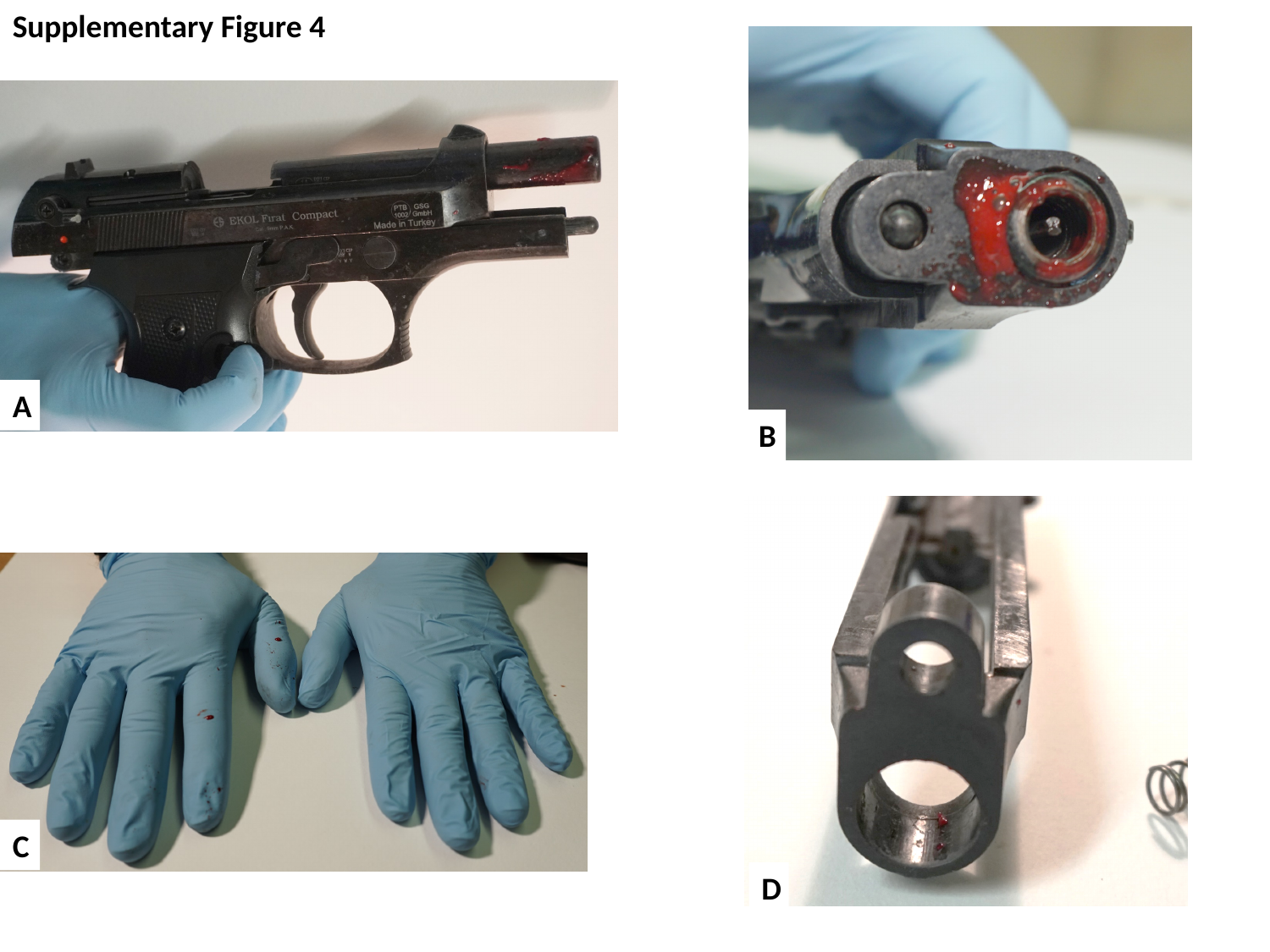

Supplementary Figure 4
A
B
C
D

### Slide 5
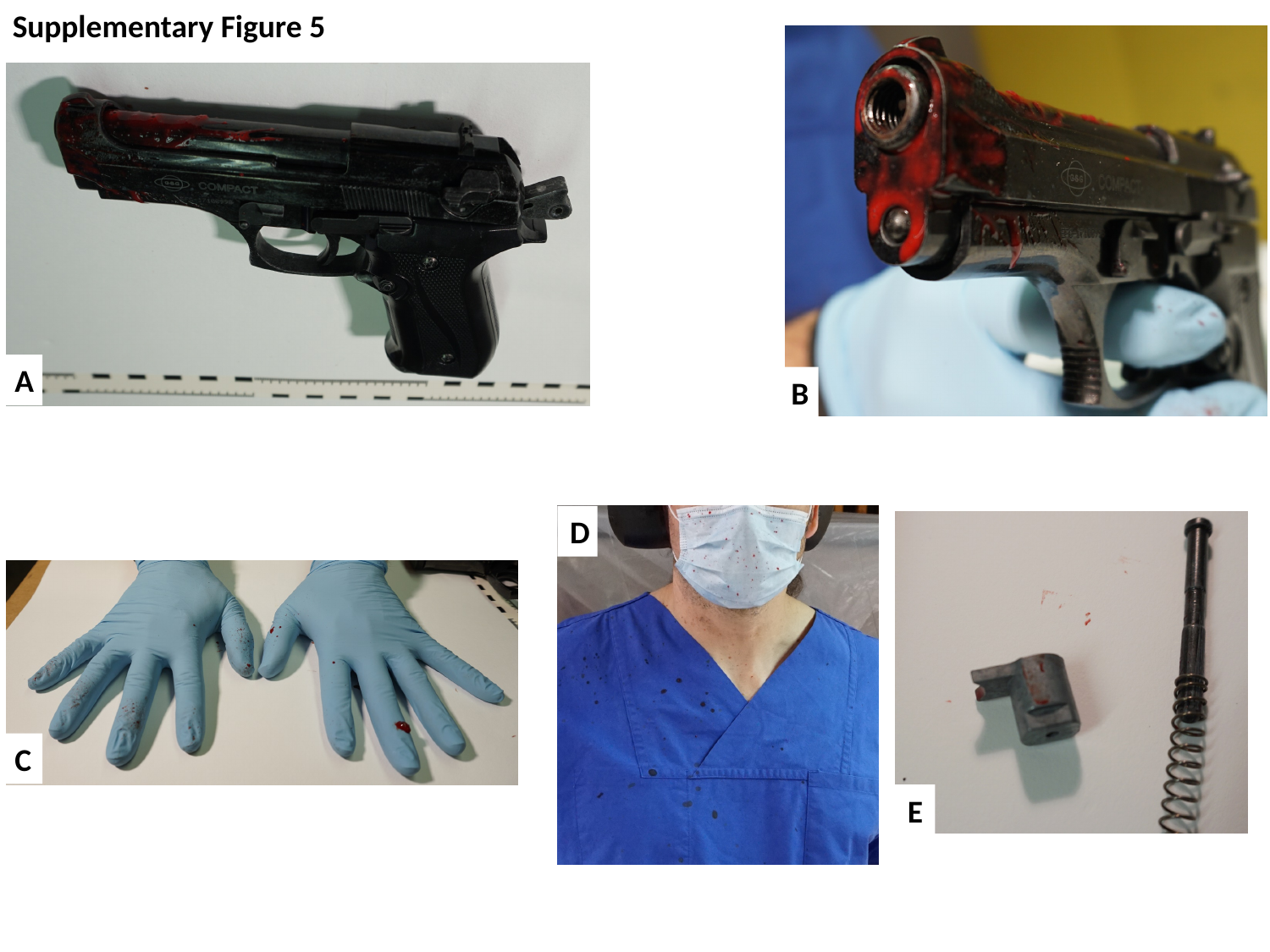

Supplementary Figure 5
A
B
D
C
E

### Slide 6
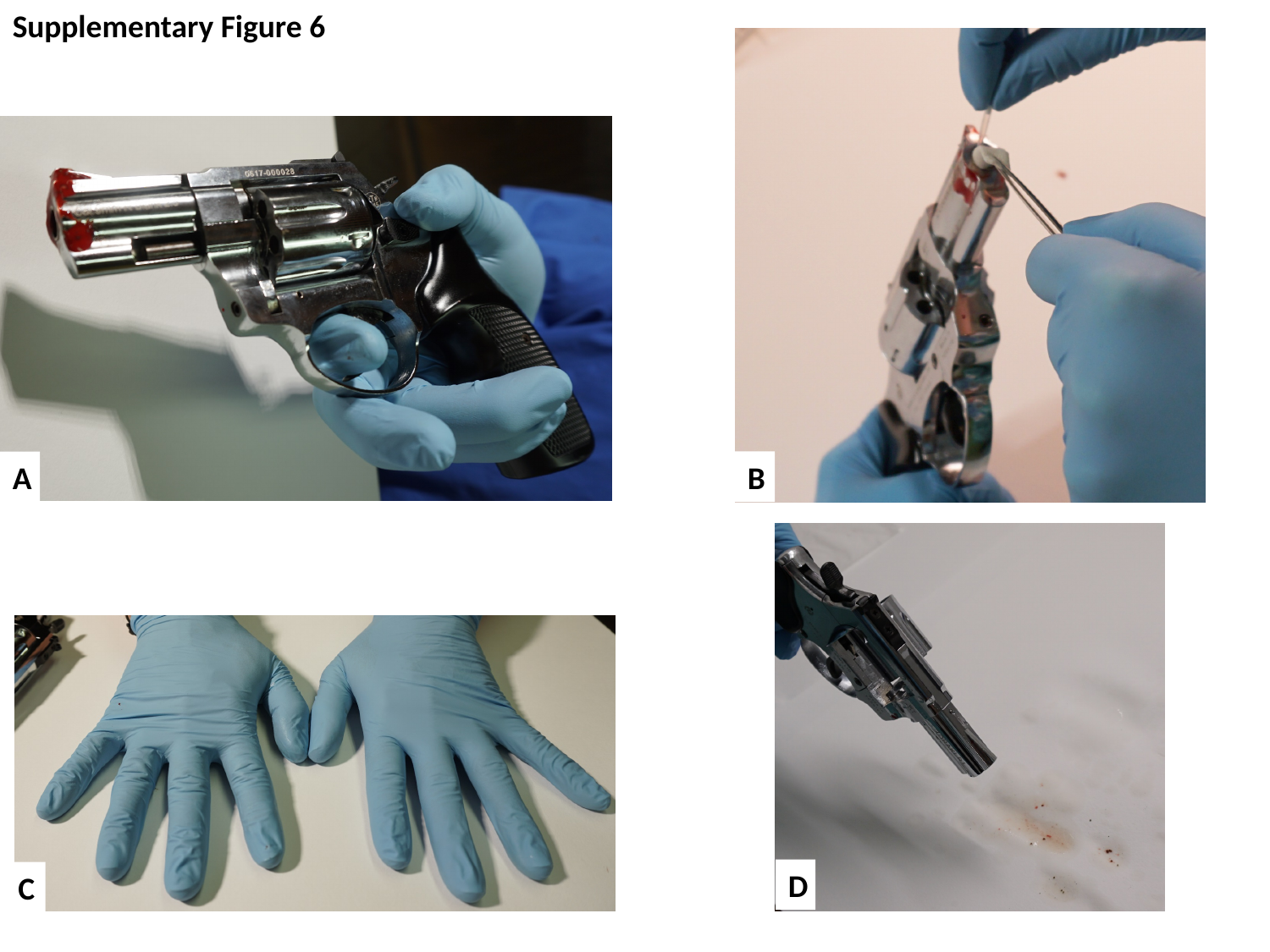

Supplementary Figure 6
B
A
D
C

### Slide 7
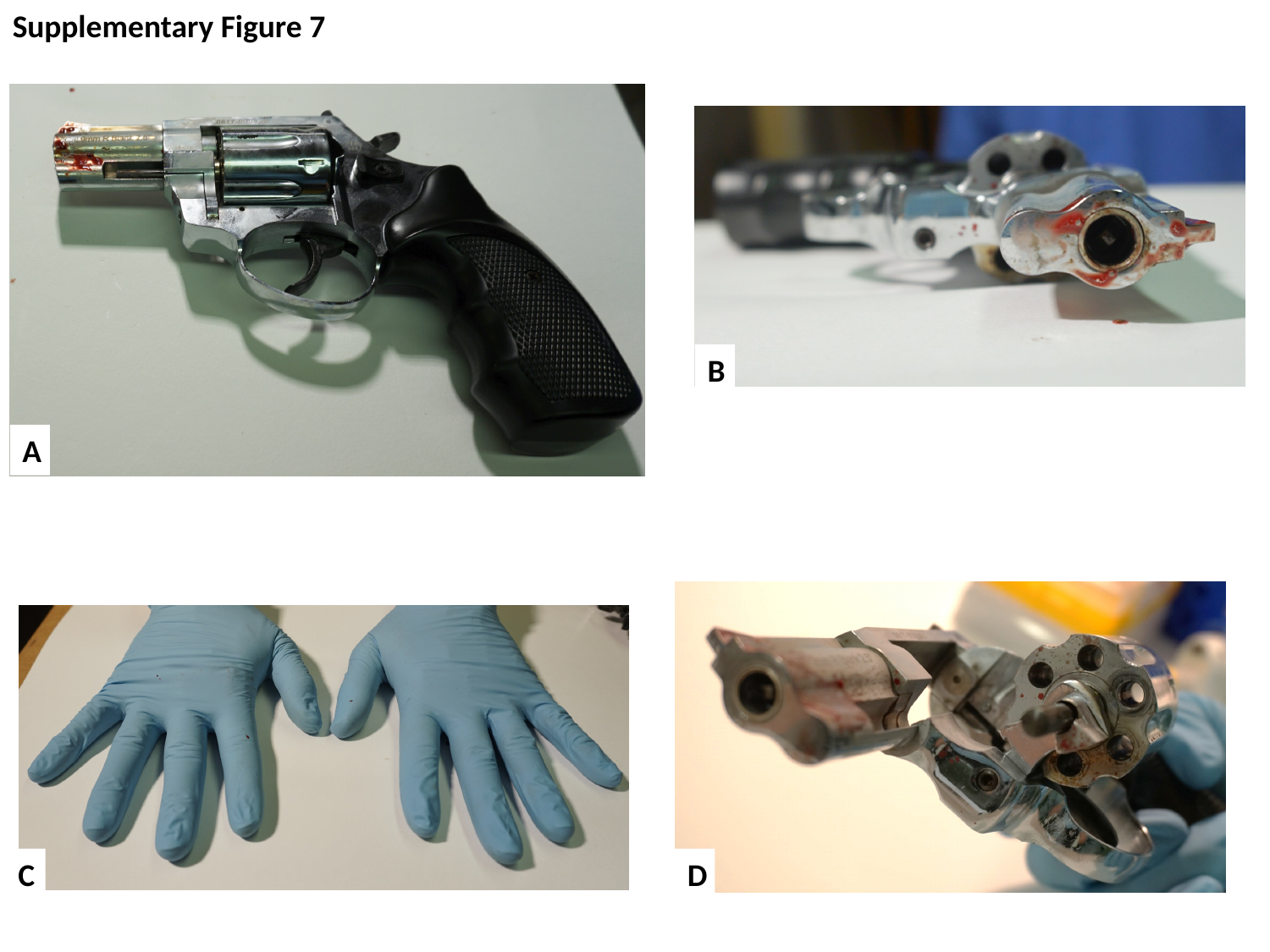

Supplementary Figure 7
B
A
C
D

### Slide 8
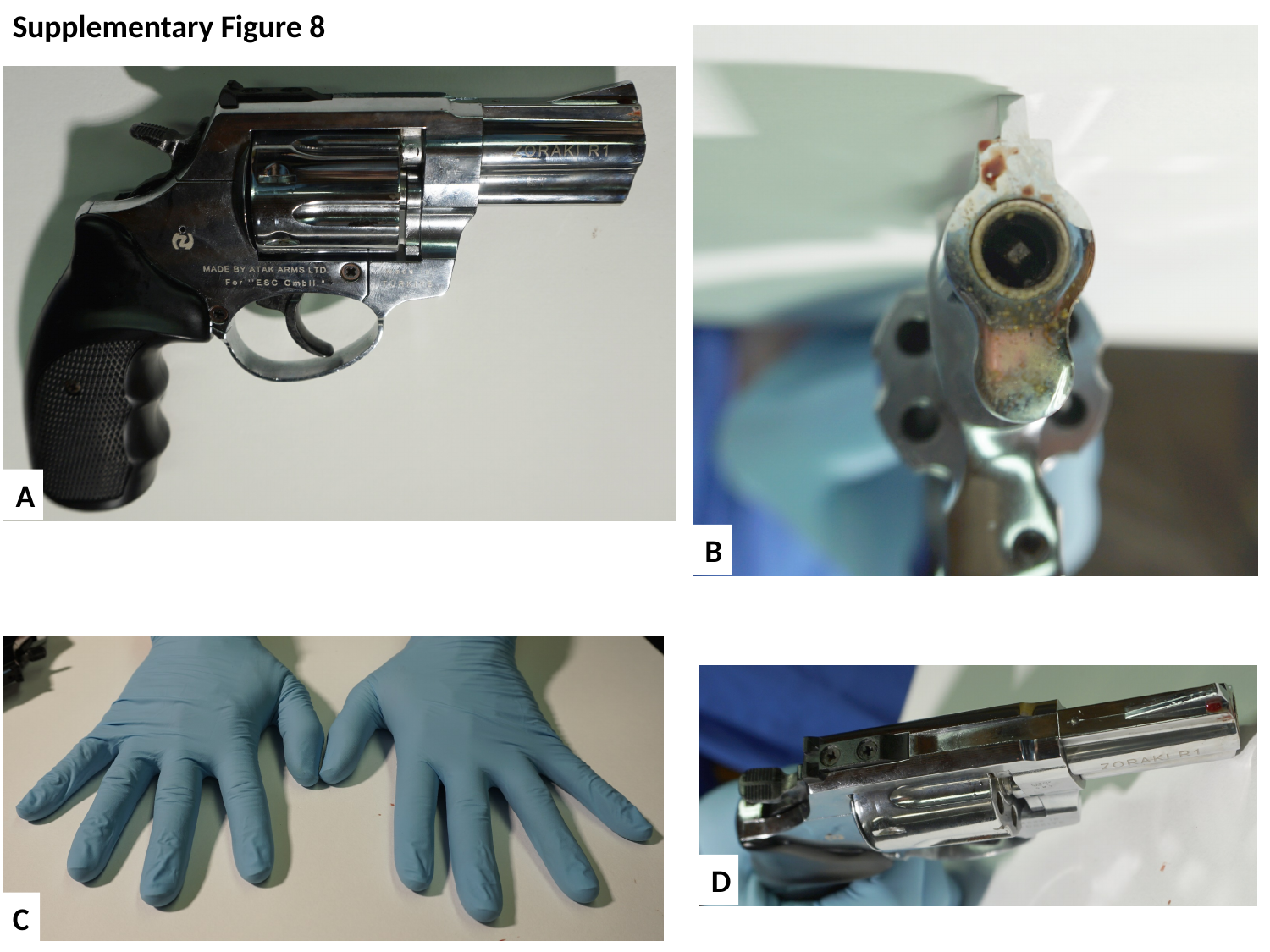

Supplementary Figure 8
A
B
D
C

### Slide 9
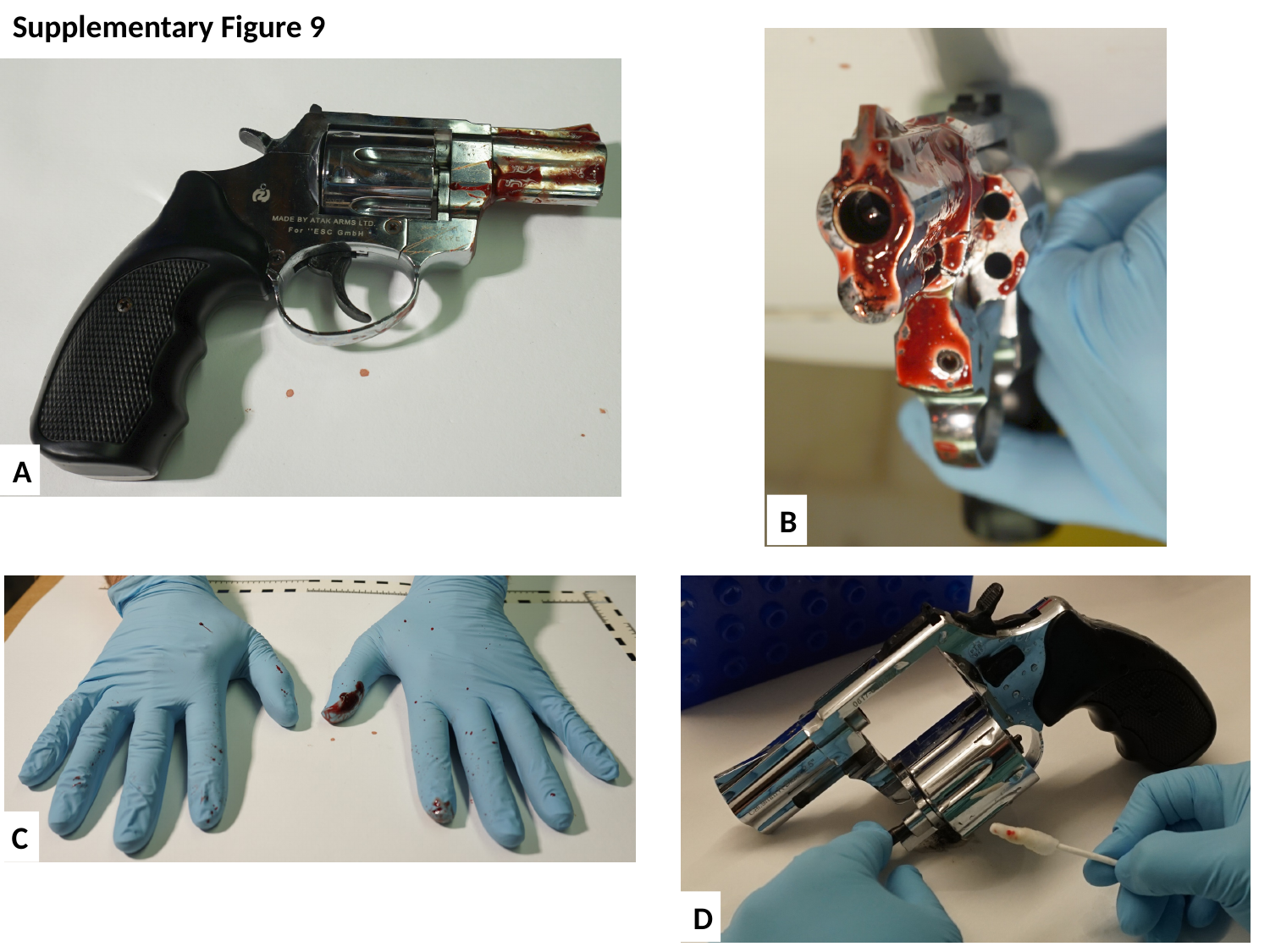

Supplementary Figure 9
A
B
C
D

### Slide 10
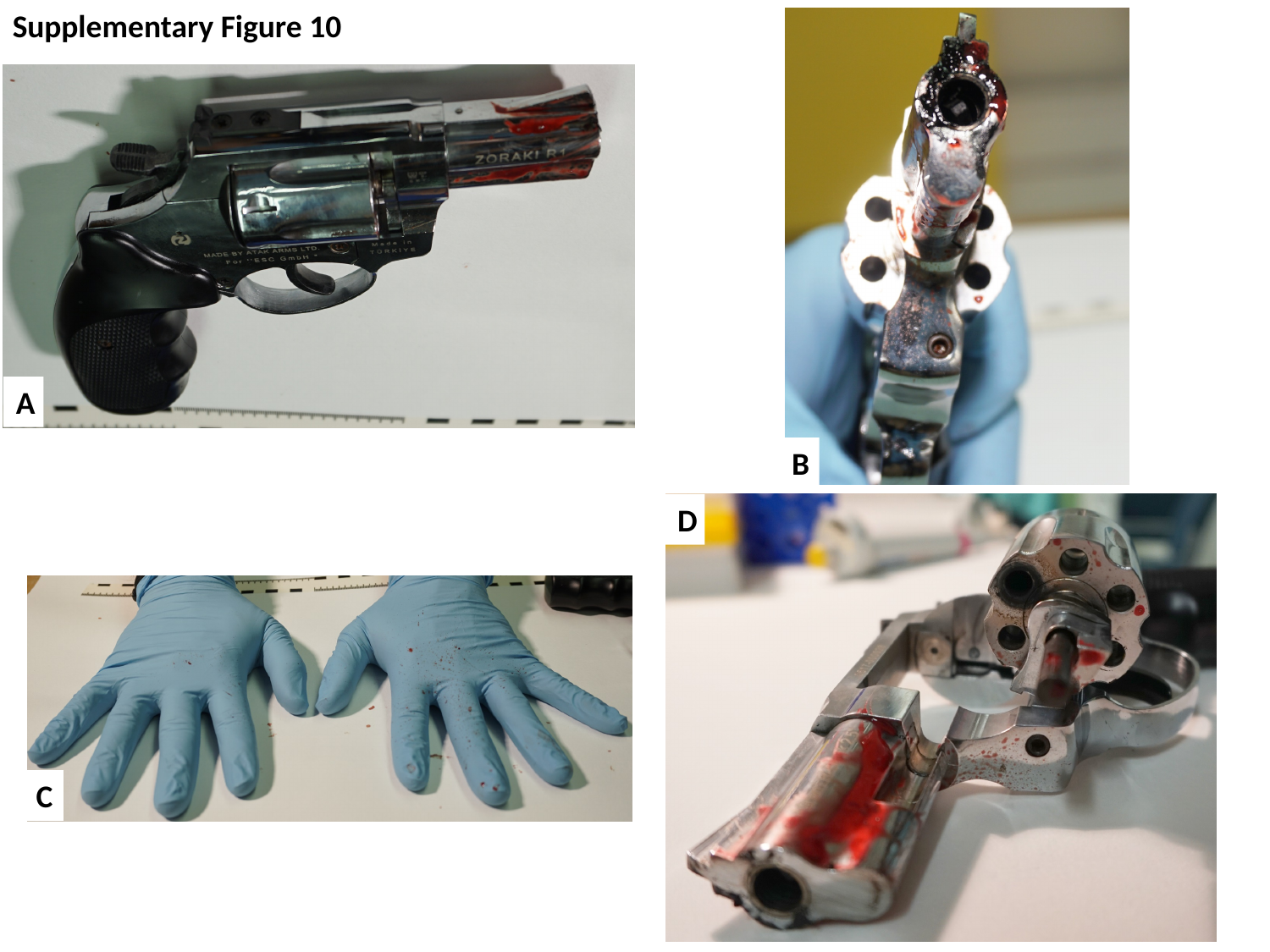

Supplementary Figure 10
A
B
D
C

### Slide 11
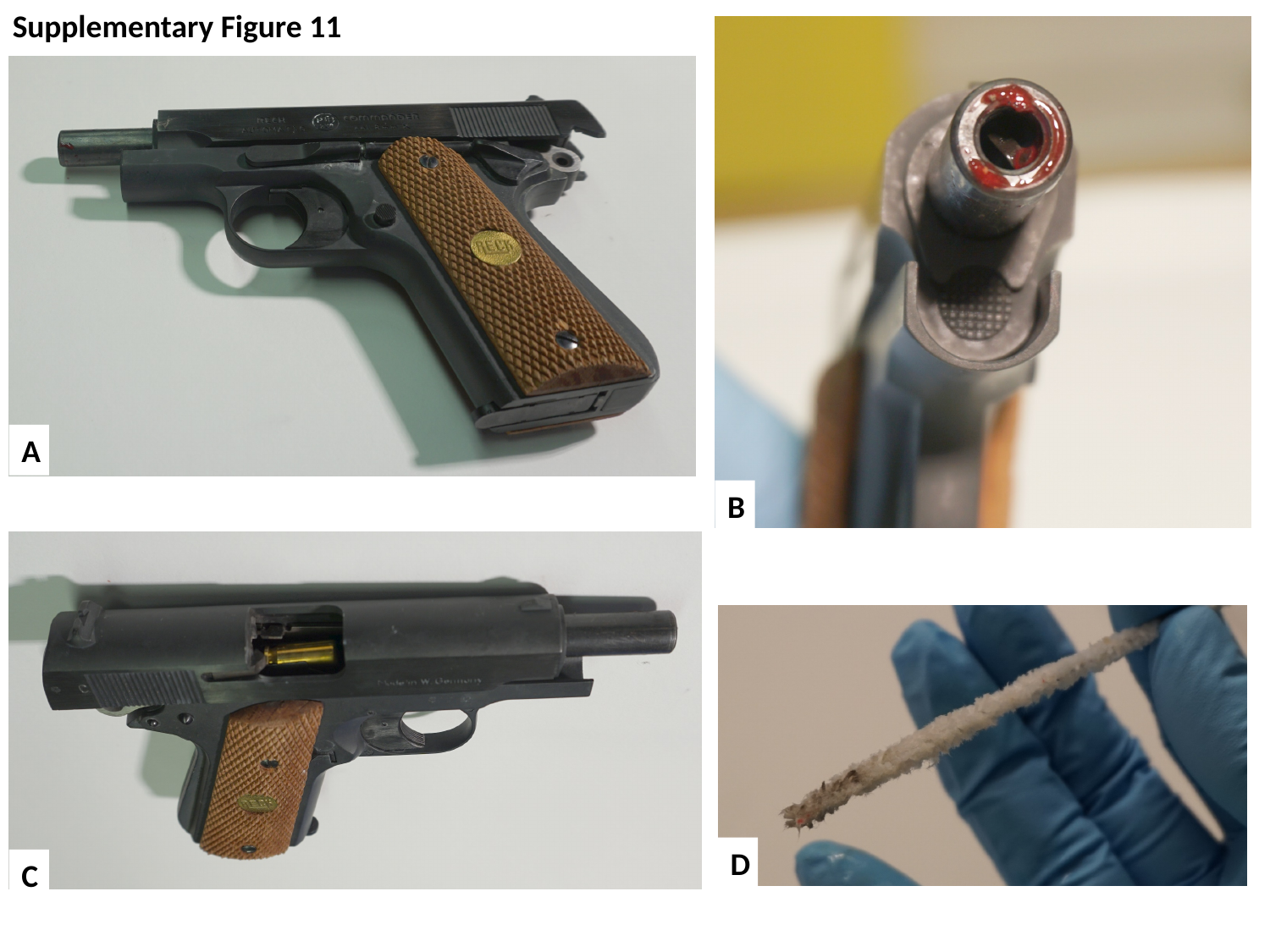

Supplementary Figure 11
A
B
D
C

### Slide 12
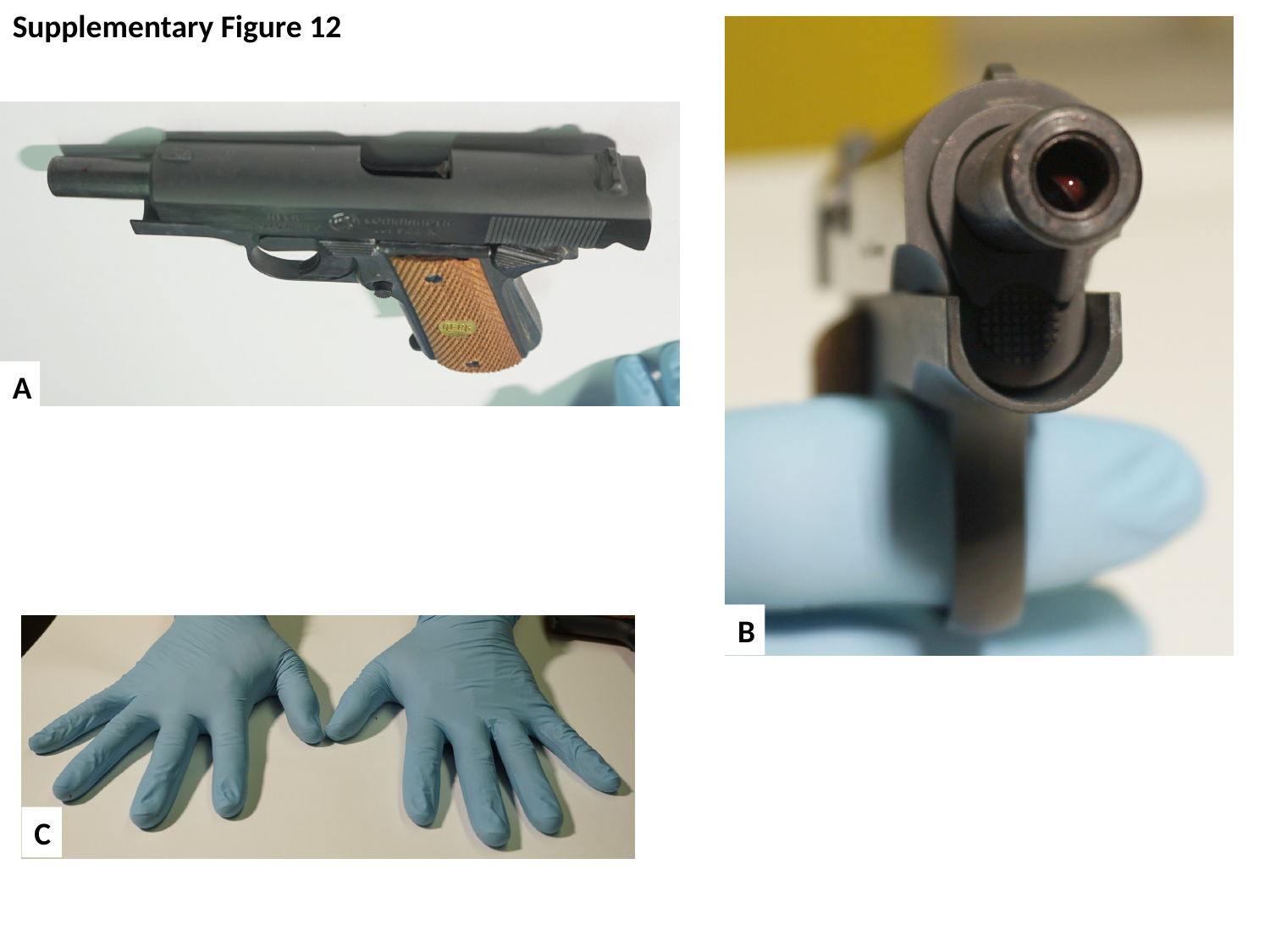

Supplementary Figure 12
A
B
C

### Slide 13
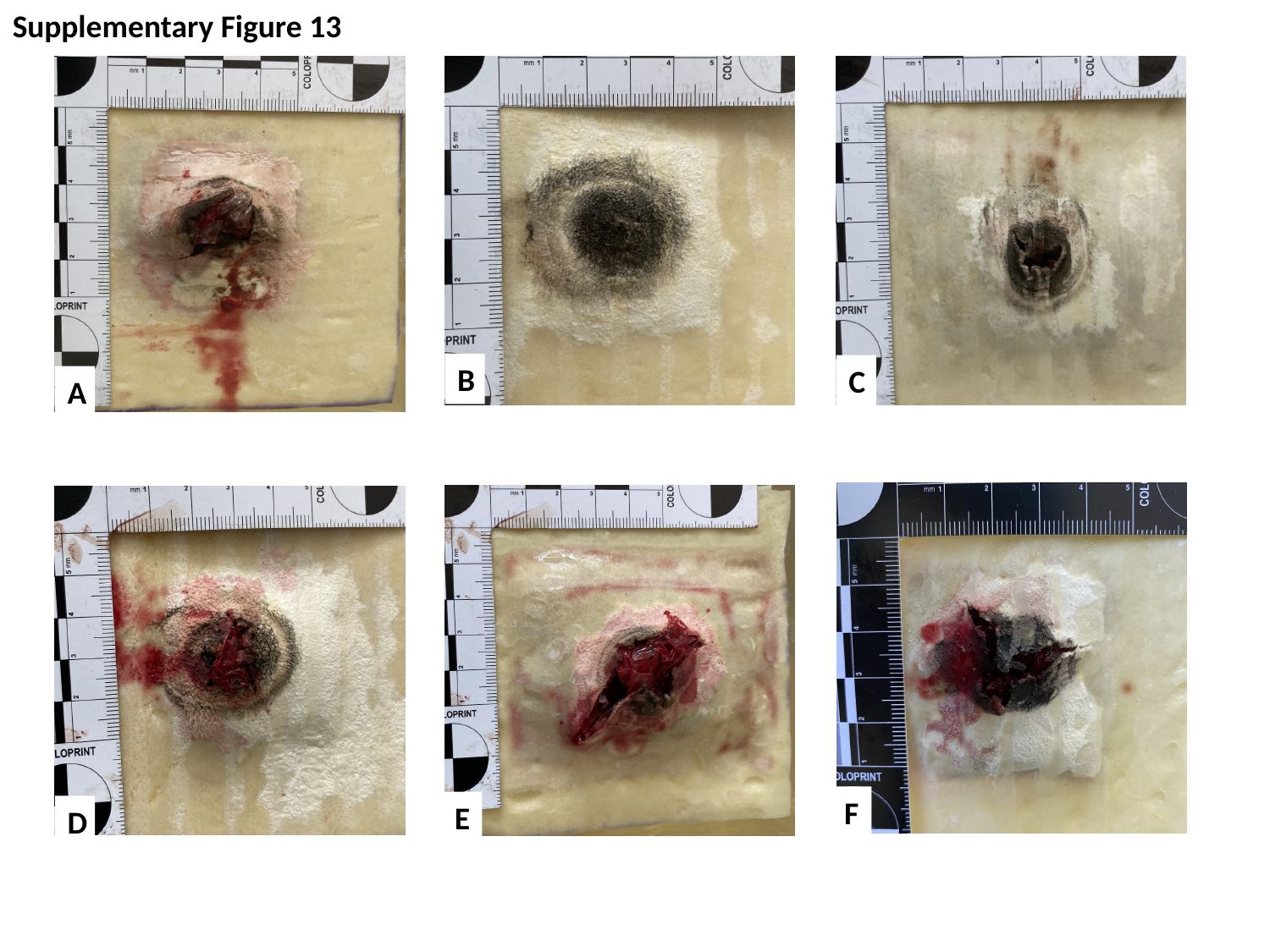

Supplementary Figure 13
B
C
A
F
E
D

### Slide 14
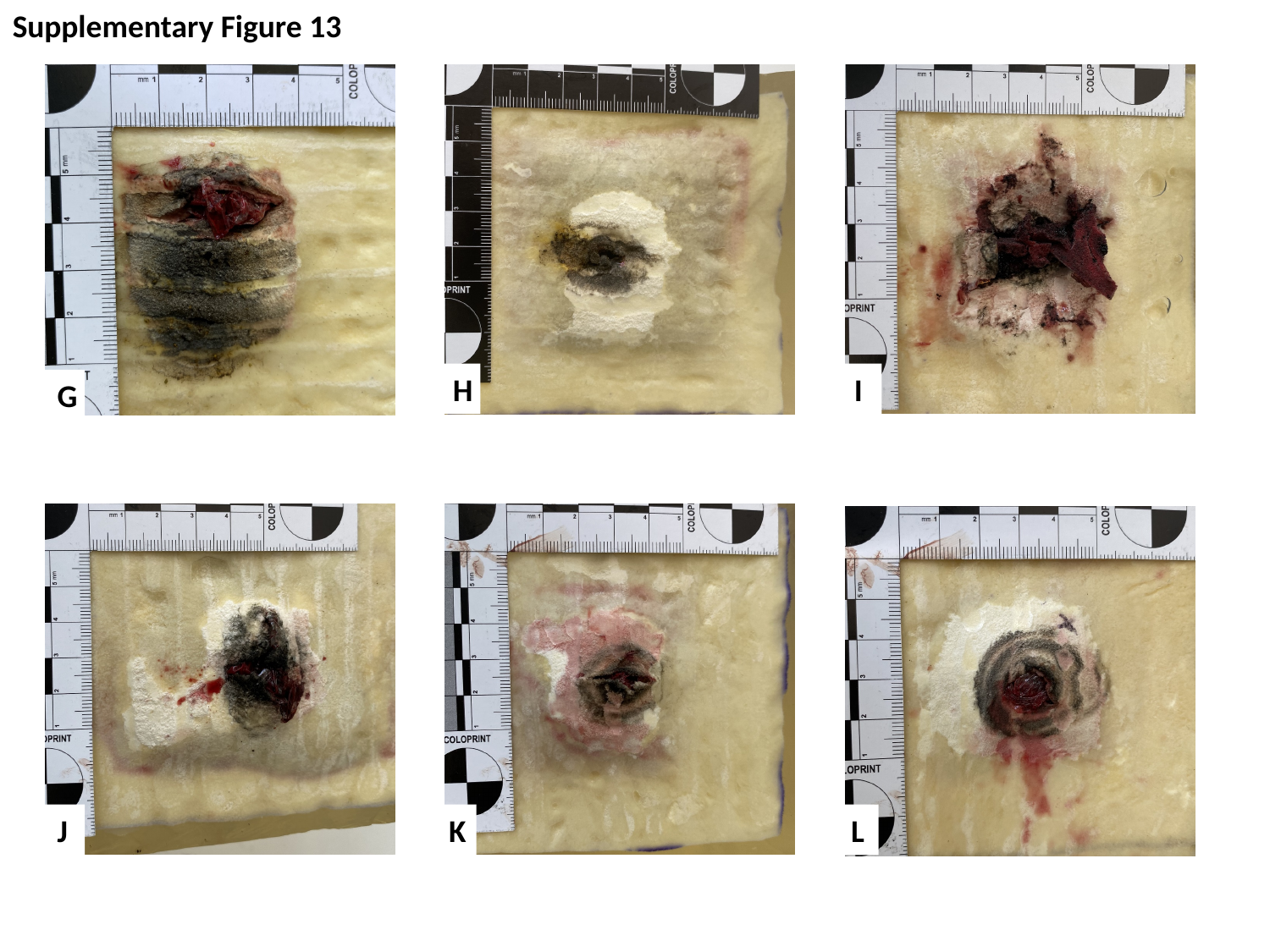

Supplementary Figure 13
H
I
G
J
K
L

### Slide 15
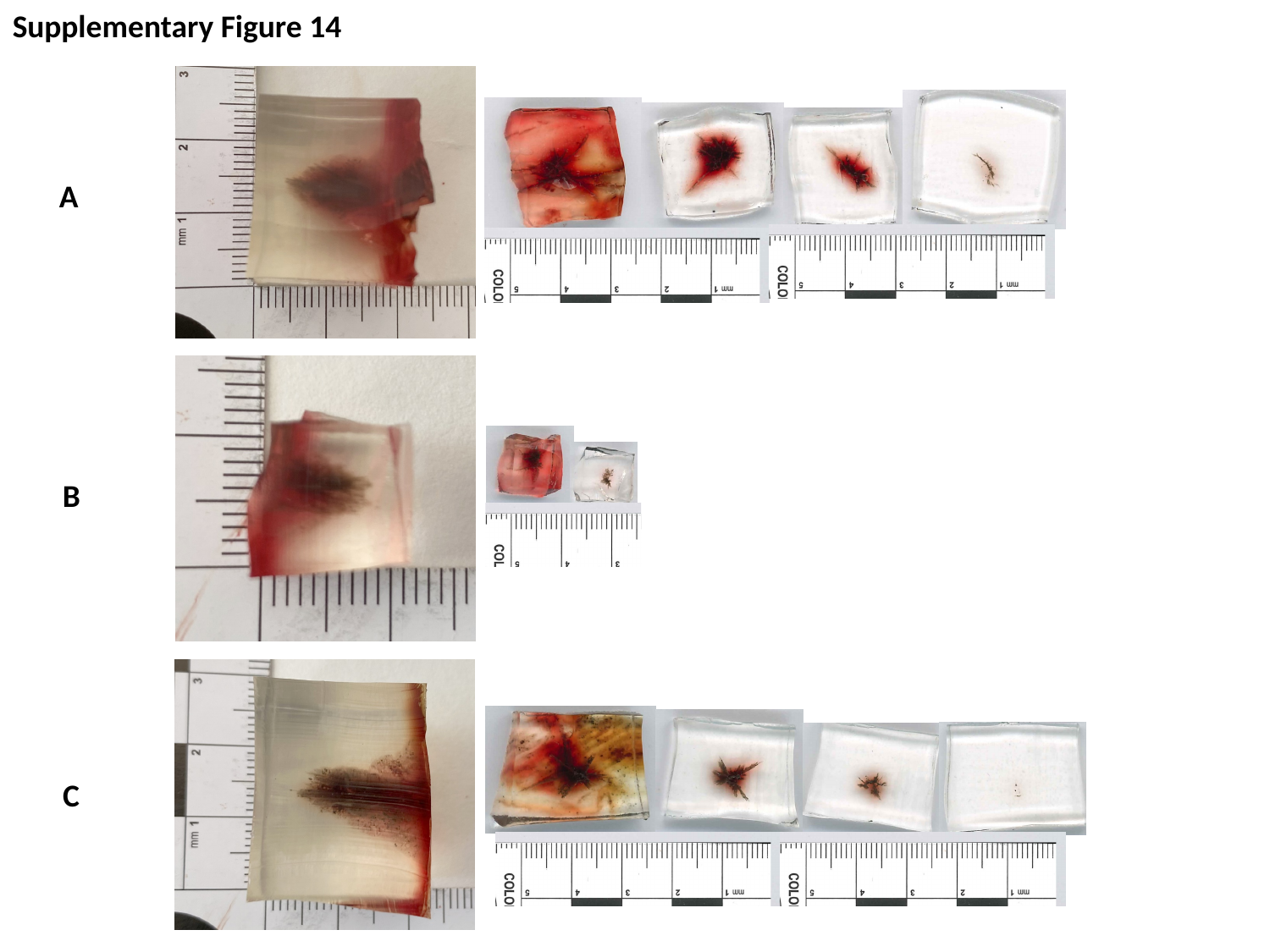

Supplementary Figure 14
A
B
C

### Slide 16
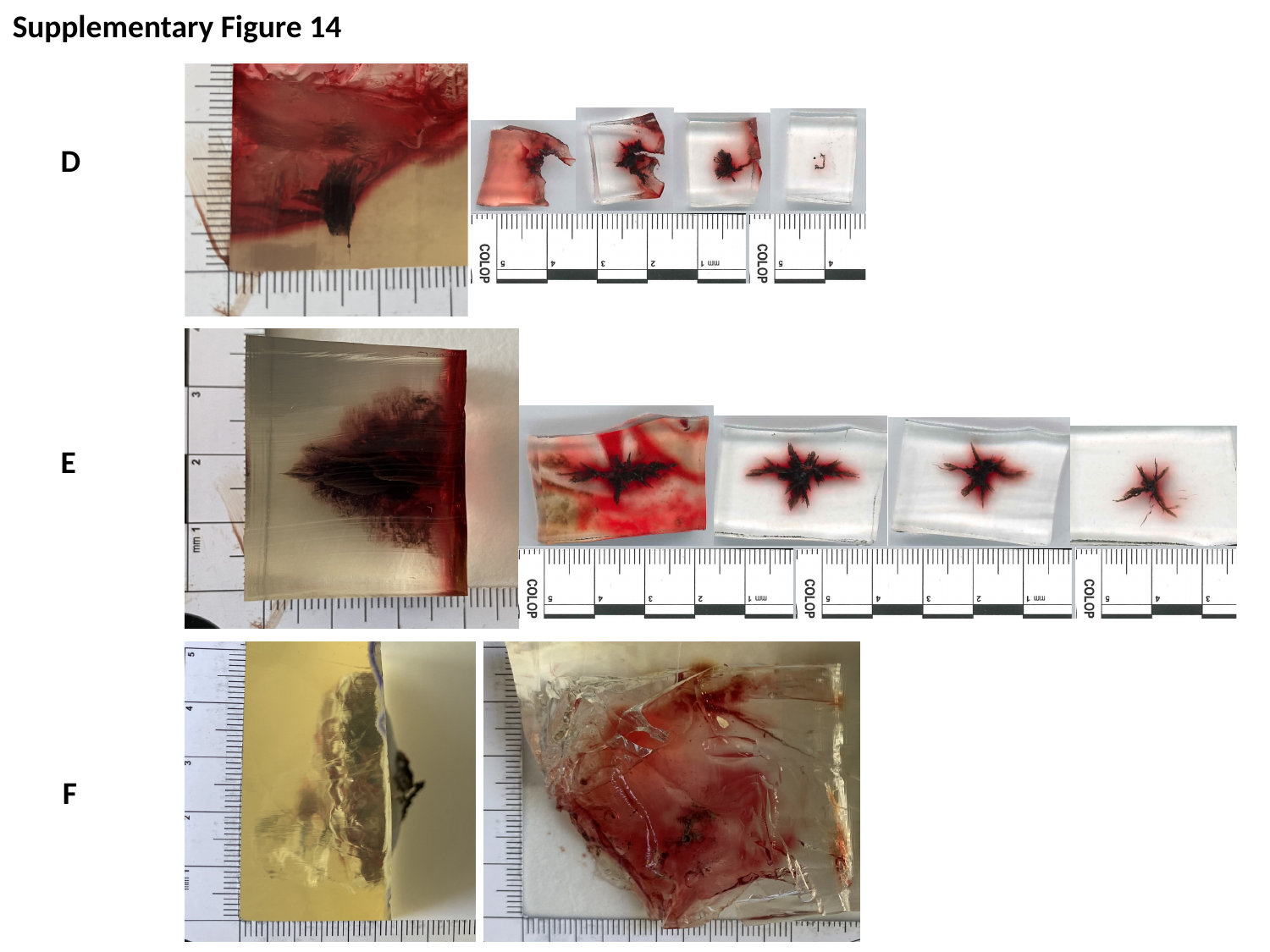

Supplementary Figure 14
D
E
F

### Slide 17
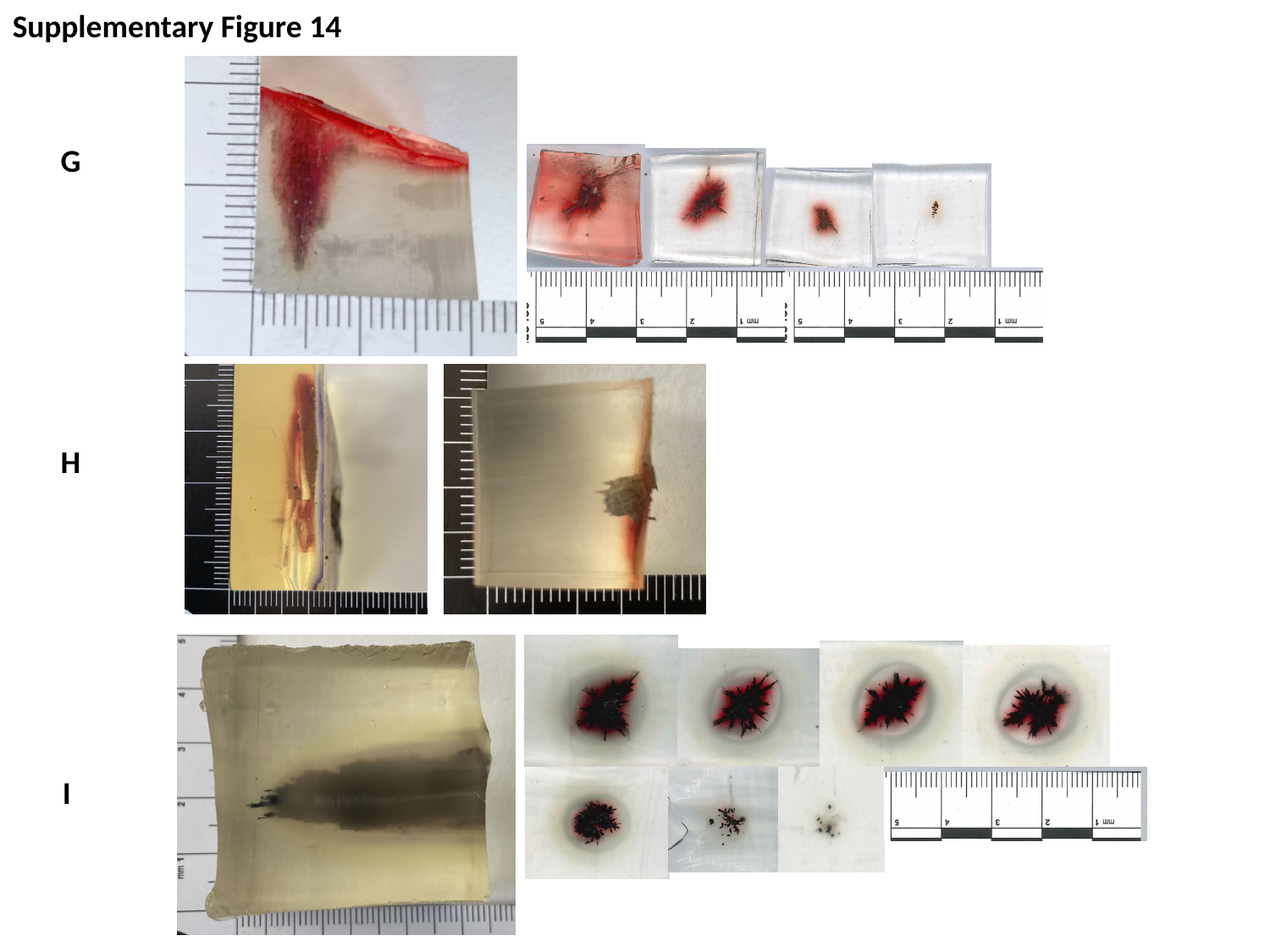

Supplementary Figure 14
G
H
I

### Slide 18
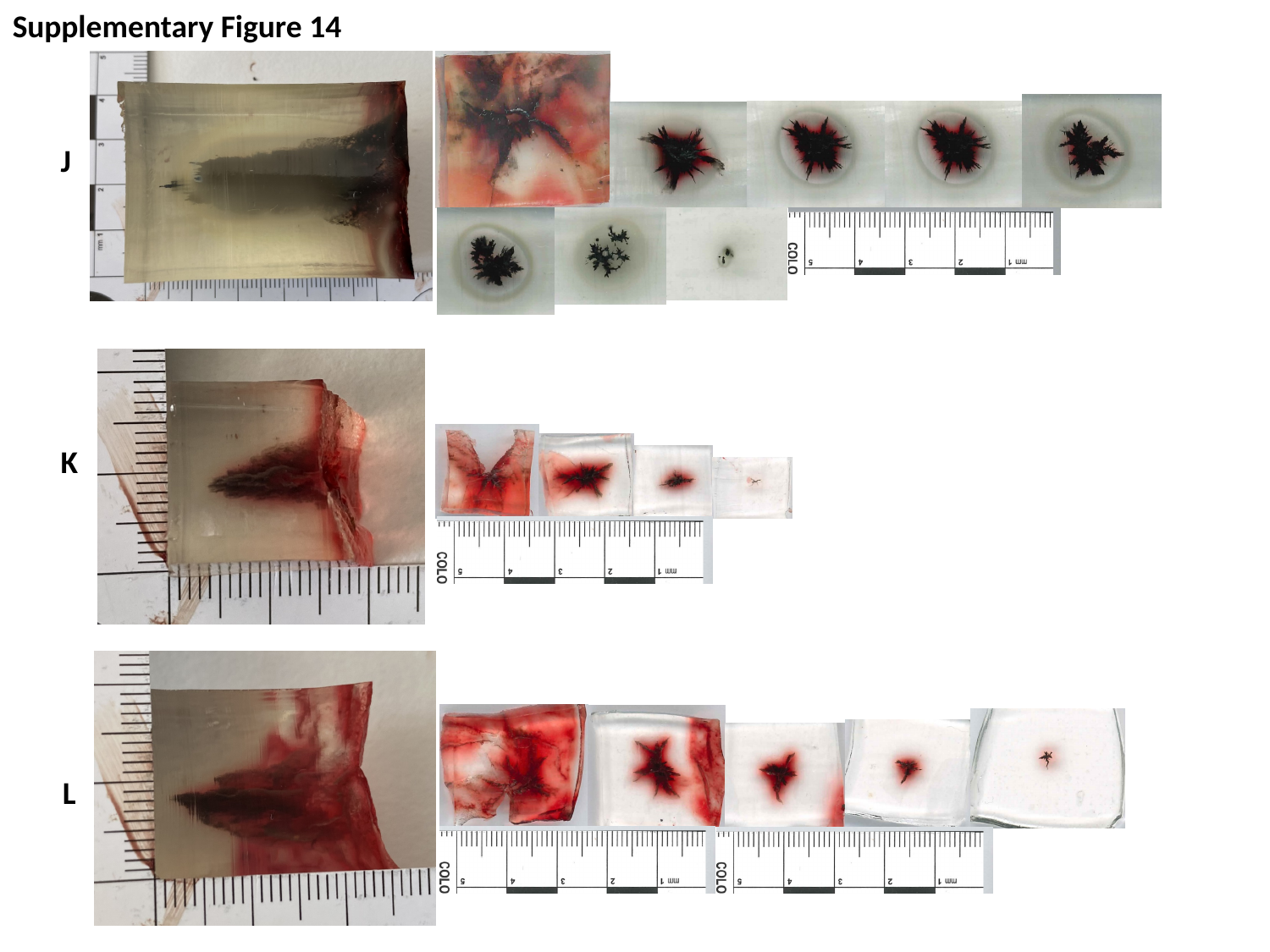

Supplementary Figure 14
J
K
L

### Slide 19
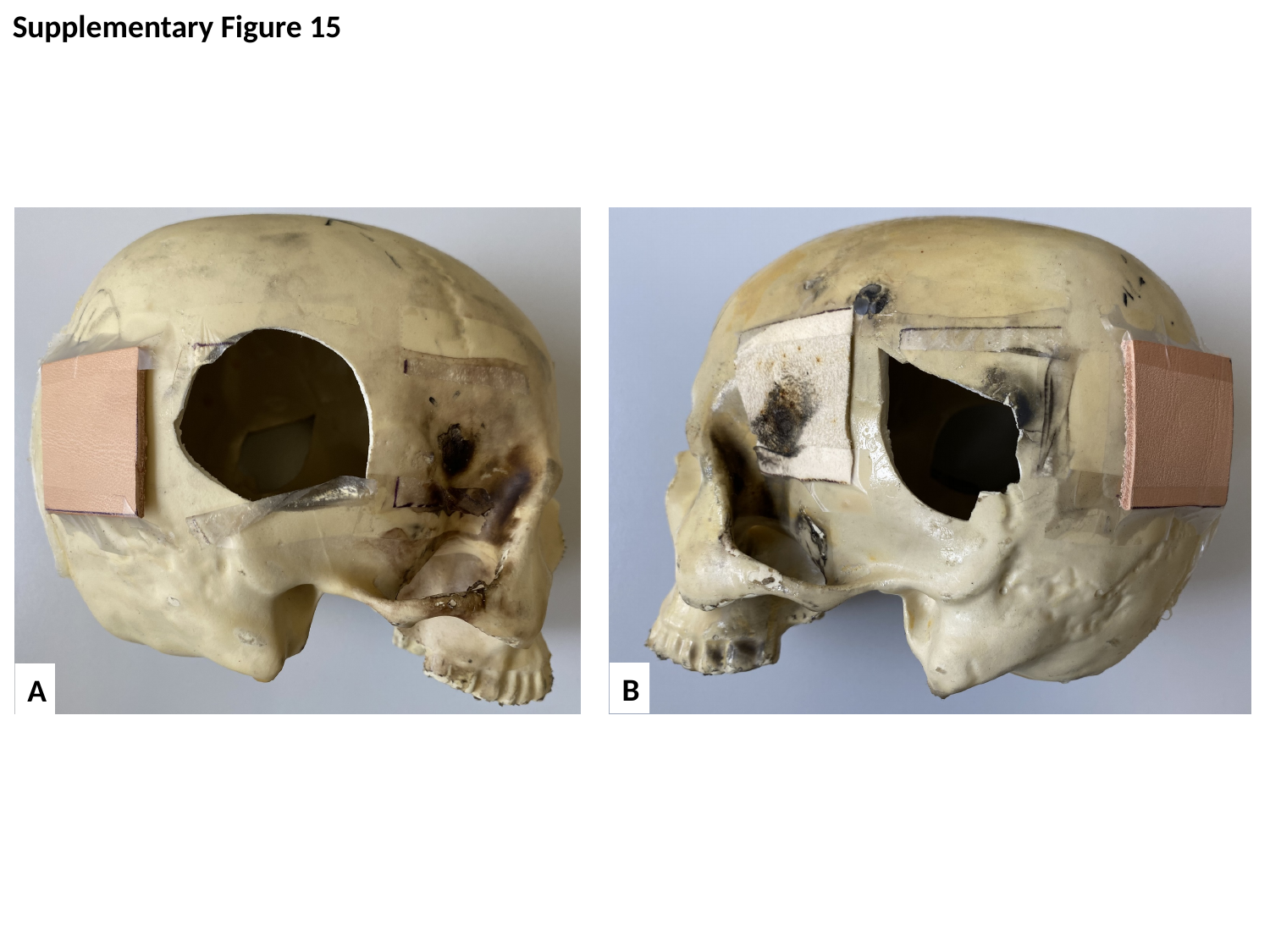

Supplementary Figure 15
B
A

### Slide 20
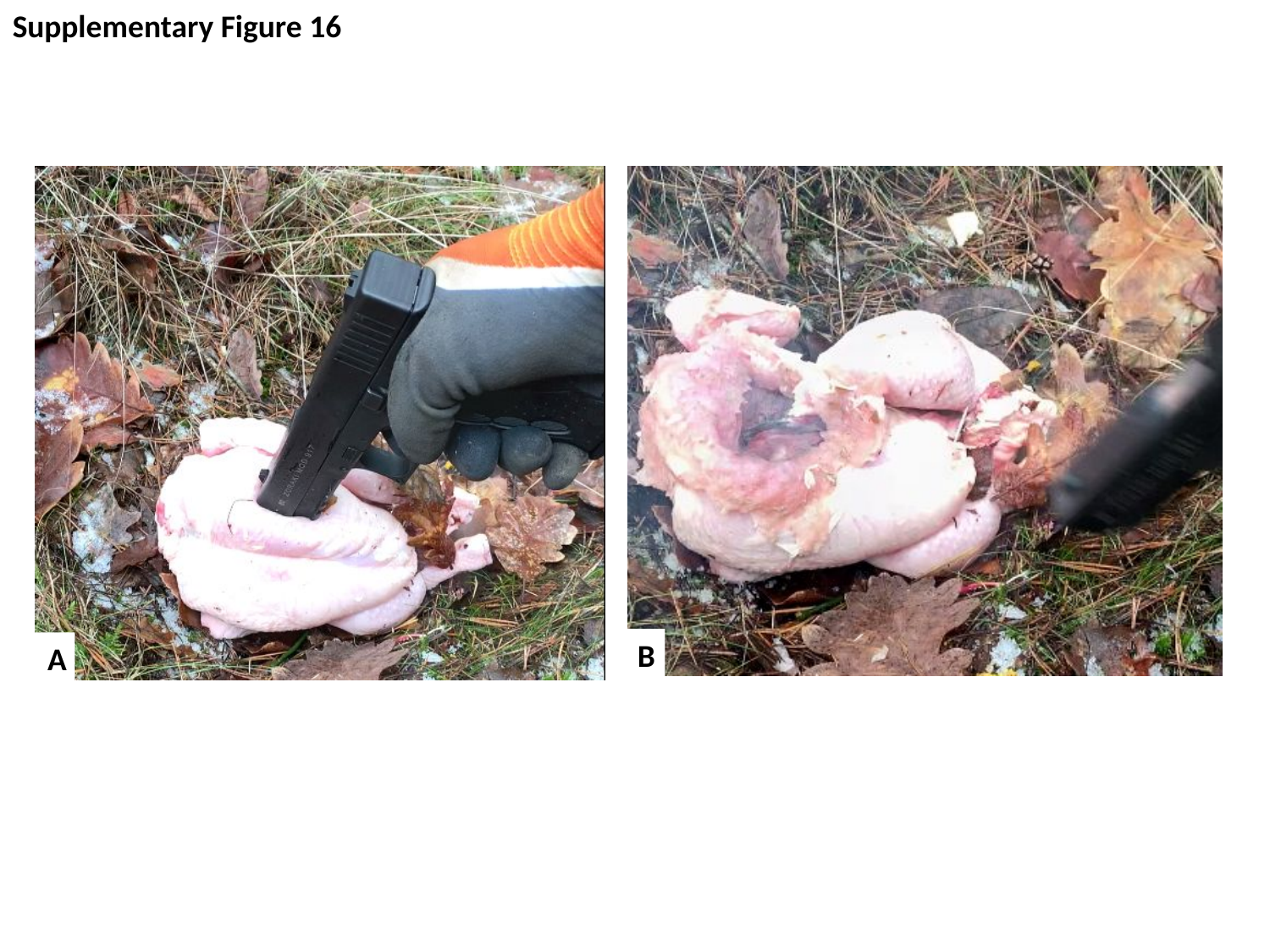

Supplementary Figure 16
B
A

### Slide 21
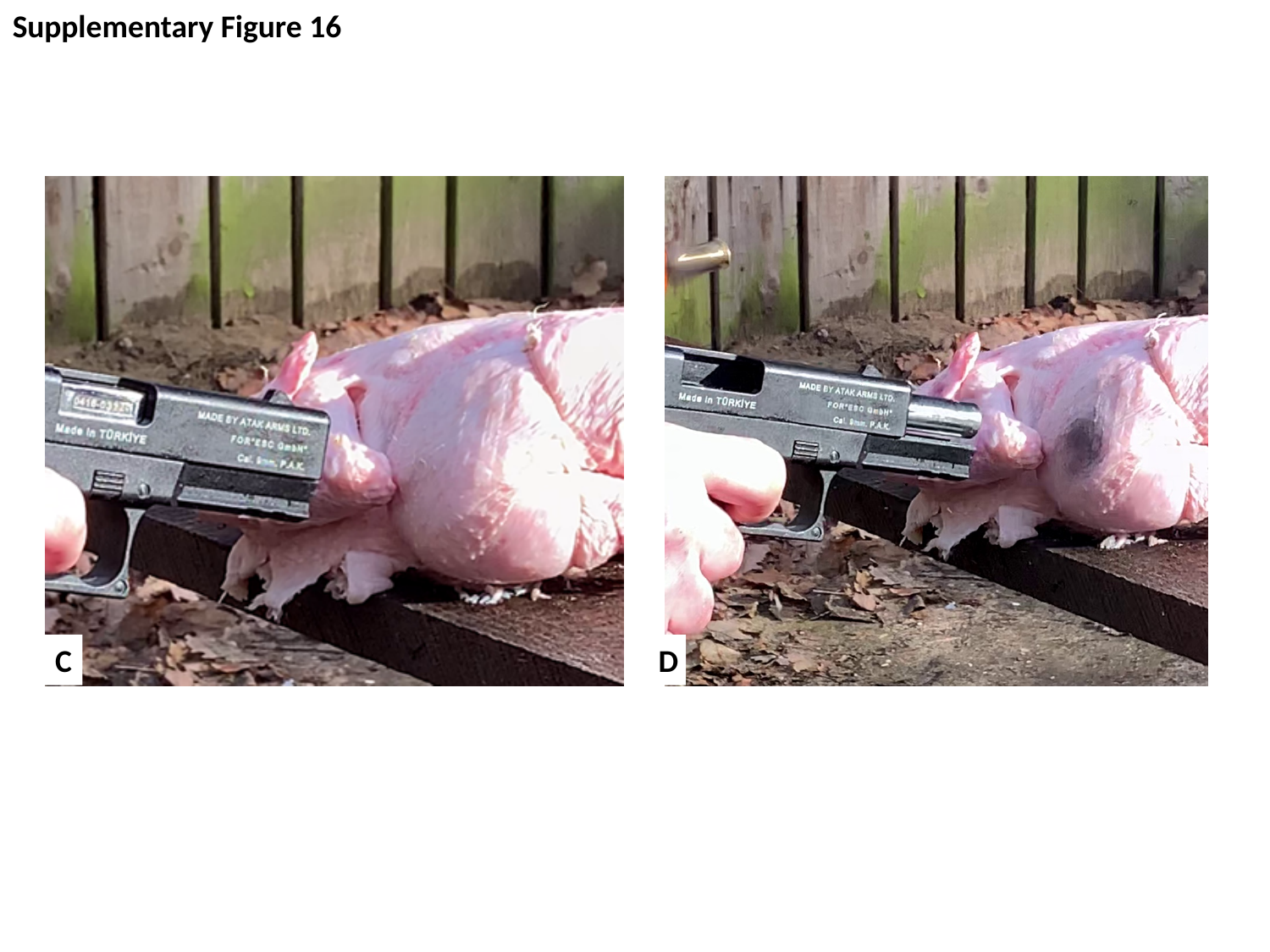

Supplementary Figure 16
C
D

### Slide 22
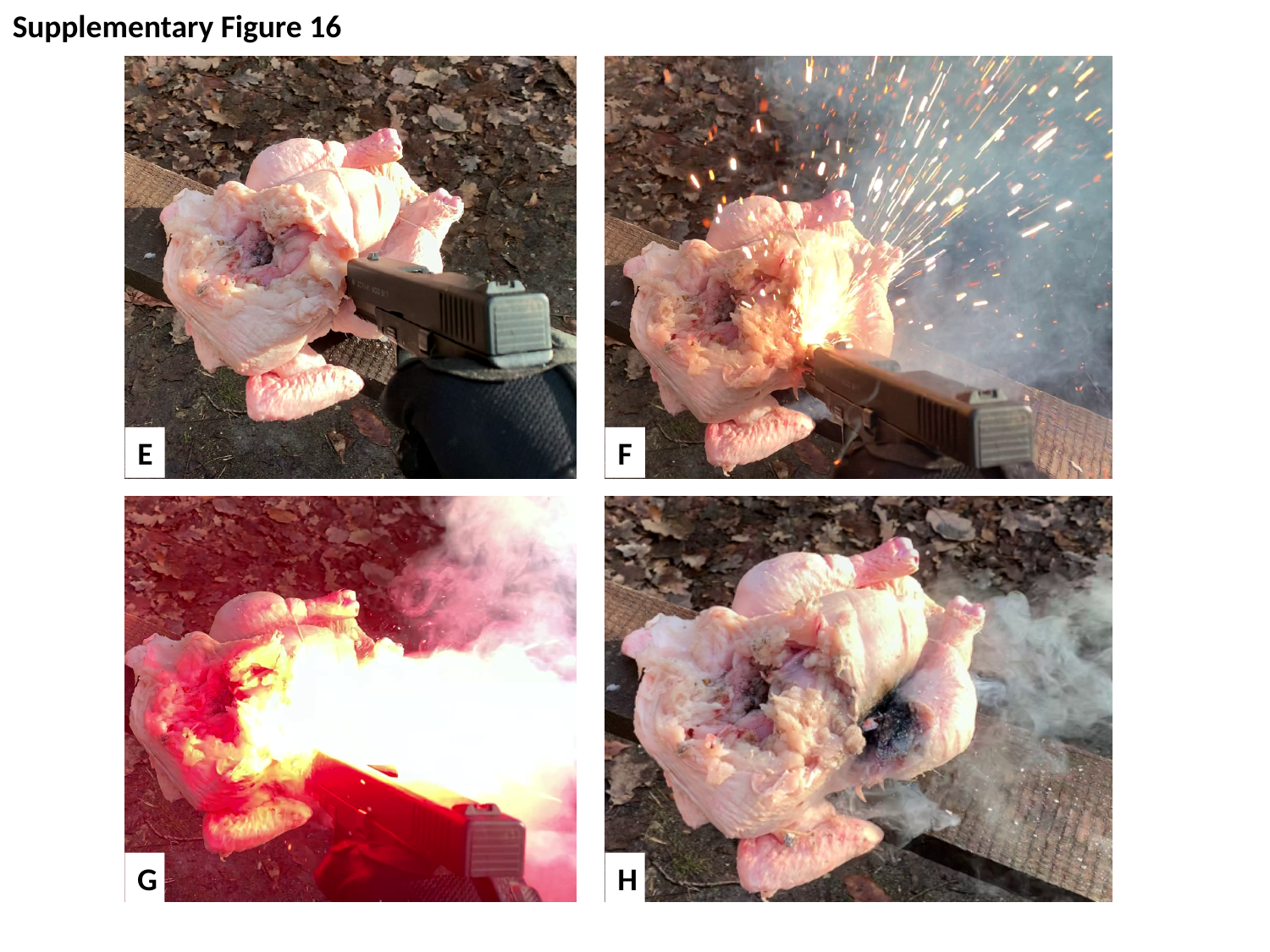

Supplementary Figure 16
E
F
G
H

### Slide 23
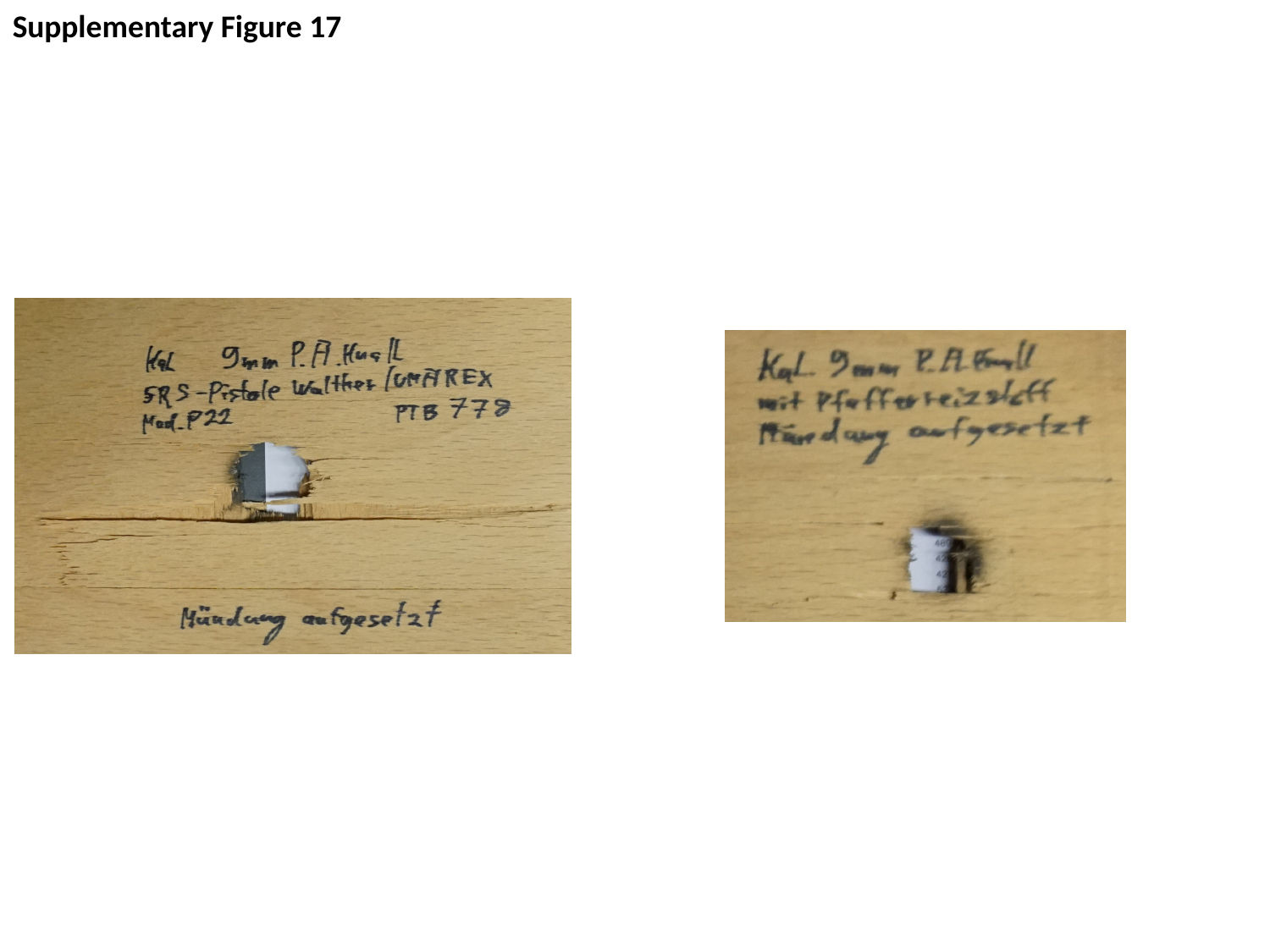

Supplementary Figure 17
